# Effectiveness of mRNA-1273 against Delta, Mu, and other emerging variants

**DOI:** 10.1101/2021.09.29.21264199

**Authors:** Katia J. Bruxvoort, Lina S. Sy, Lei Qian, Bradley K. Ackerson, Yi Luo, Gina S. Lee, Yun Tian, Ana Florea, Michael Aragones, Julia E. Tubert, Harpreet S. Takhar, Jennifer H. Ku, Yamuna D. Paila, Carla A. Talarico, Hung Fu Tseng

## Abstract

**Background:** Real-world studies have found high vaccine effectiveness (VE) of mRNA-based COVID-19 vaccines, but reduced VE against the Delta variant and waning protection have been reported, with few studies examining mRNA-1273 variant-specific VE.

**Methods:** We conducted a test-negative case-control study at Kaiser Permanente Southern California. Whole genome sequencing was conducted for SARS-CoV-2 positive specimens collected from 3/1/2021 to 7/27/2021. Test-positive cases were matched 1:5 to test-negative controls on age, sex, race/ethnicity, and specimen collection date. Outcomes included SARS-CoV-2 infection and hospitalization. Exposures were 2 doses or 1 dose of mRNA-1273 ≥14 days prior to specimen collection versus no COVID-19 vaccination. Conditional logistic regression was used to compare odds of vaccination among cases versus controls, adjusting for confounders. VE was calculated as (1-odds ratio)x100%.

**Results:** The study included 8,153 cases and their matched controls. Two-dose VE (95% confidence interval) was 86.7% (84.3-88.7%) against Delta infection, 98.4% (96.9-99.1%) against Alpha, 90.4% (73.9-96.5%) against Mu, 96-98% against other identified variants, and 79.9% (76.9-82.5%) against unidentified variants. VE against Delta declined from 94.1% (90.5-96.3%) 14-60 days after vaccination to 80.0% (70.2-86.6%) 151-180 days after vaccination. Waning was less pronounced for non-Delta variants. VE against Delta was lower among individuals aged ≥65 years (75.2% [59.6-84.8%]) than those aged 18-64 years (87.9% [85.5-89.9%]). VE against Delta hospitalization was 97.6% (92.8-99.2%). One-dose VE was 77.0% (60.7-86.5%) against Delta infection.

**Conclusions:** Two doses of mRNA-1273 were highly effective against all SARS-CoV-2 variants. However, VE against Delta moderately declined with increasing time since vaccination.

**Trial Registration Number:** Not applicable

**Funding:** Moderna Inc.

## INTRODUCTION

Vaccines to prevent coronavirus disease 2019 (COVID-19) were developed rapidly in response to the COVID-19 pandemic. In clinical trials, mRNA-based COVID-19 vaccines, mRNA-1273 (Moderna Inc, Cambridge, USA) and BNT162b2 (Pfizer Inc, New York, USA; BioNTech Manufacturing GmbH, Mainz, Germany) were highly efficacious (94% and 95%, respectively) against symptomatic COVID-19.^1,2^ After receiving emergency use authorization in the United States (US) in December 2020,^3,4^ these vaccines were deployed in phased mass vaccination programs for high risk and general populations.

Subsequently, multiple real-world studies conducted before the SARS-CoV-2 Delta variant (B.1.617.2 and AY lineage) became predominant reported high vaccine effectiveness (VE) of mRNA-based vaccines against COVID-19 infection (e.g., 82%-100%)^5-8^ and COVID-19 hospitalization (e.g., 87-96%).^9,10^ Few of these studies identified variant-specific VE. In a study in Qatar, VE of mRNA-1273 against infection with Alpha (B.1.1.7) and Beta (B.1.351) SARS-CoV-2 variants was 100% and 96.4%, respectively.^11^ A study in Canada found VE of mRNA-based vaccines against Alpha and Beta/Gamma (P.1) infection of 90% and 88%, respectively.^12^

However, as Delta became predominant, concerns arose that mRNA-based vaccines could be less effective.^13,14^ The higher transmissibility of Delta led to a surge in infections, hospitalizations, and deaths in the US.^15^ These cases have occurred overwhelmingly among unvaccinated individuals but have also included breakthrough cases.^16,17^ Although studies have found sustained VE of mRNA-based vaccines against COVID-19 hospitalization during periods overlapping with or during the Delta surge,^18-20^ decreased VE against infection with Delta in some studies has been reported (e.g., 51-75%).^20-22^ It is unclear if these findings are due to lower protection against Delta, waning vaccine immunity over time, or other factors.

Furthermore, few studies have examined VE specifically for mRNA-1273 against Delta or other SARS-CoV-2 variants. Such studies are critically needed to inform ongoing decisions around booster doses and development of vaccines that may offer broad protection against SARS-CoV-2 variants. Thus, we evaluated the VE of mRNA-1273 against variants including Delta by time since vaccination at Kaiser Permanente Southern California (KPSC).

## METHODS

### Study setting

KPSC is an integrated health care system with 15 hospitals and associated medical offices across Southern California. The population of over 4.6 million members with diverse sociodemographic characteristics is generally representative of the underlying population.^23^ Comprehensive electronic health records (EHR) provide detailed data on all aspects of clinical care, including diagnoses, laboratory tests, procedures, and pharmacy records. The KPSC Institutional Review Board approved this study.

KPSC began administering COVID-19 vaccines on December 18, 2020, following state guidelines for vaccine prioritization.^24^ COVID-19 vaccinations received outside KPSC are regularly imported into the EHR from external sources, including the California Immunization Registry (CAIR),^25^ CalVax (Cal Poly Pomona mass vaccination site), Care Everywhere (system on the Epic EHR platform that allows different health care systems to exchange patient medical information), claims (e.g., retail pharmacies), and member self-report (with valid documentation).

### Laboratory methods

Molecular diagnostic testing for SARS-CoV-2 is widely available at KPSC for symptomatic and asymptomatic individuals and is required prior to procedures or hospital admission. Specimens are primarily collected using nasopharyngeal/oropharyngeal swabs or saliva (asymptomatic individuals only) and tested using the RT-PCR TaqPath™ COVID-19 High-Throughput Combo Kit (Thermo Fisher Scientific, California, USA). Beginning in March 2021, KPSC began sending all positive SARS-CoV-2 specimens from both symptomatic and asymptomatic individuals, regardless of cycle threshold (Ct) values, to a commercial laboratory (Helix, California, USA) for whole genome sequencing (WGS), as described in **Supplementary Methods**.

### Study design

The study used a test-negative design to examine VE of mRNA-1273 against SARS-CoV-2 variants. Individuals who had a SARS-CoV-2 positive test sent for WGS or a negative test from March 1, 2021 to July 27, 2021 were eligible for inclusion in the study if they were age ≥18 years and had ≥12 months of KPSC membership as of the specimen collection date. Individuals were excluded if they received a COVID-19 vaccine other than mRNA-1273, received 2 doses of mRNA-1273 <24 days apart or <14 days prior to specimen collection date, received >2 doses of mRNA-1273 prior to the specimen collection date, or had a positive SARS-CoV-2 test or COVID-19 diagnosis code between 12/18/2020 and 2/28/2021 or ≤90 days prior to positive test date. Separate analyses were conducted for each SARS-CoV-2 variant, selected based on scientific relevance and prevalence in the KPSC population. These included Delta (B.1.617.2, AY.*), Alpha (B.1.1.7), Epsilon (B.1.427, B.1.429), Gamma (P.1, P.1.1, P.1.2), Iota (B.1.526, B.1.526.1, B.1.526.2), Mu (B.1.621, B.1.621.1), and other (Beta, Eta, Kappa, and any other variants). Test-positive cases were defined as the first positive SARS-CoV-2 specimen identified by WGS. Cases for which WGS failed were examined as a separate category (“unidentified variants”). COVID-19 hospitalized test-positive cases were defined as a variant with specimen collection date ≤7 days before or during COVID-19 hospitalization confirmed by chart review. Test-negative controls were selected from eligible individuals with a negative SARS-CoV-2 test. Cases and controls were matched 1:5 on age (18–44 years, 45–64 years, 65–74 years, and ≥75 years), sex, race/ethnicity (Non-Hispanic White, Non-Hispanic Black, Hispanic, Non-Hispanic Asian, and Other/Unknown), and specimen collection date (±10 days). The exposure of interest was receipt of 2 doses administered ≥24 days apart or 1 dose of mRNA-1273 ≥14 days prior to specimen collection date.

Demographic and clinical covariates were extracted from EHR (**Table S1**). Variables assessed at specimen collection date included socioeconomic status (Medicaid, neighborhood median household income), medical center area, pregnancy status, and KPSC physician/employee status. Variables assessed in the 2 years prior to specimen collection date included smoking and body mass index (BMI). Variables assessed in the year prior to specimen collection date included Charlson comorbidity score, autoimmune conditions, health care utilization (virtual, outpatient, emergency department, and inpatient encounters), preventive care (other vaccinations, screenings, and well-visits), chronic diseases (kidney disease, heart disease, lung disease, liver disease, diabetes), and frailty index. Other variables included history of SARS-CoV-2 molecular test performed from March 1, 2020 to specimen collection date (irrespective of result), history of COVID-19 diagnosis (SARS-CoV-2 positive molecular test or a COVID-19 diagnosis code) from March 1, 2020 to specimen collection date, and immunocompromised status.

### Statistical analyses

The distribution of variants was described by vaccination status and by calendar time. Characteristics of cases and controls for each analysis were described and compared using the χ^2^ test or Fisher’s exact test for categorical variables and the two-sample t-test or Wilcoxon rank-sum test for continuous variables. Conditional logistic regression was used to estimate the odds ratios (OR) and 95% confidence interval (95% CI) for vaccination, comparing cases and controls. Analyses were adjusted for potential confounders, determined by absolute standardized differences (ASD) >0.1 and p-value <0.1, or scientific relevance. VE was calculated as (1 – adjusted OR) x 100%. Variants with at least 20 cases were selected for analyses according to power calculations (**Supplementary Methods**).

Analyses of VE by time since receipt of second dose of mRNA-1273 (14-60 days, 61-90 days, 91-120 days, 121-150 days, 151-180 days, and >180 days) were conducted for Delta (overall and by age), non-Delta variants, and unidentified variants. All analyses were conducted using SAS software version 9.4 (Cary, USA).

## RESULTS

The study included 8,153 test-positive cases, with variants identified for 5,186 cases (63.6% overall, of which 39.4% were Delta, 27.7% Alpha, 11.4% Epsilon, 6.9% Gamma, 2.2% Iota, 1.4% Mu, and 11.1% Other) (**Table 1 and Figures S1-2**). Among fully vaccinated cases, 85.0% of identified variants were Delta. Approximately 36% of specimens failed WGS. Compared to successfully sequenced specimens, specimens that failed sequencing were more often from fully vaccinated cases (11.0% vs. 5.3%), collected via saliva from asymptomatic individuals (9.3% vs. 3.3%), and had Ct values >27 (65.5% vs. 13.7%), suggesting that low viral load coupled with the limits of detection of current molecular assays contributed to sequence failures. Additionally, some specimens that failed sequencing exhibited S gene target failure and positive results for the two other gene targets, suggesting that some unidentified variants may belong to the Alpha lineage.^26-28^

**Table 1.**
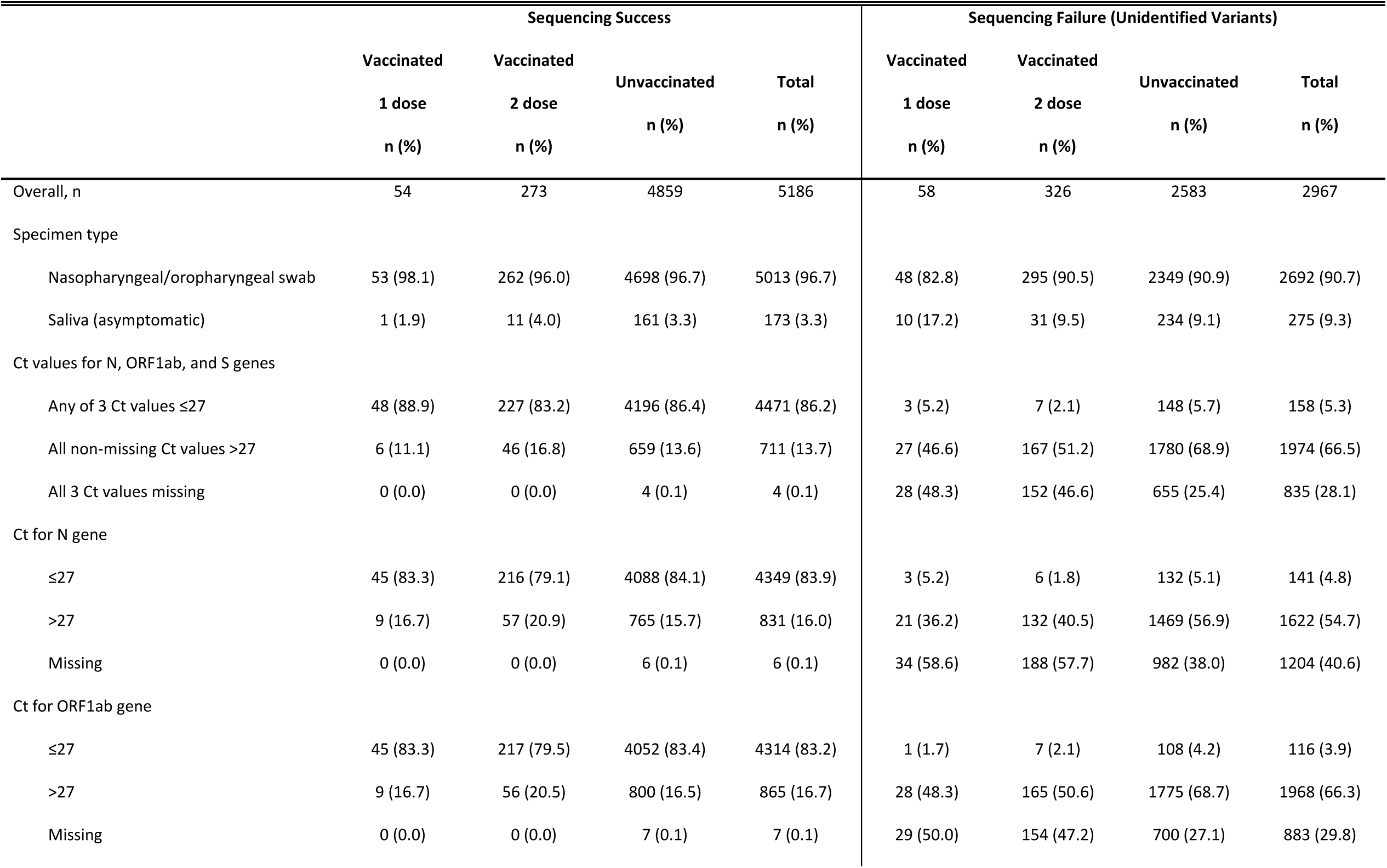
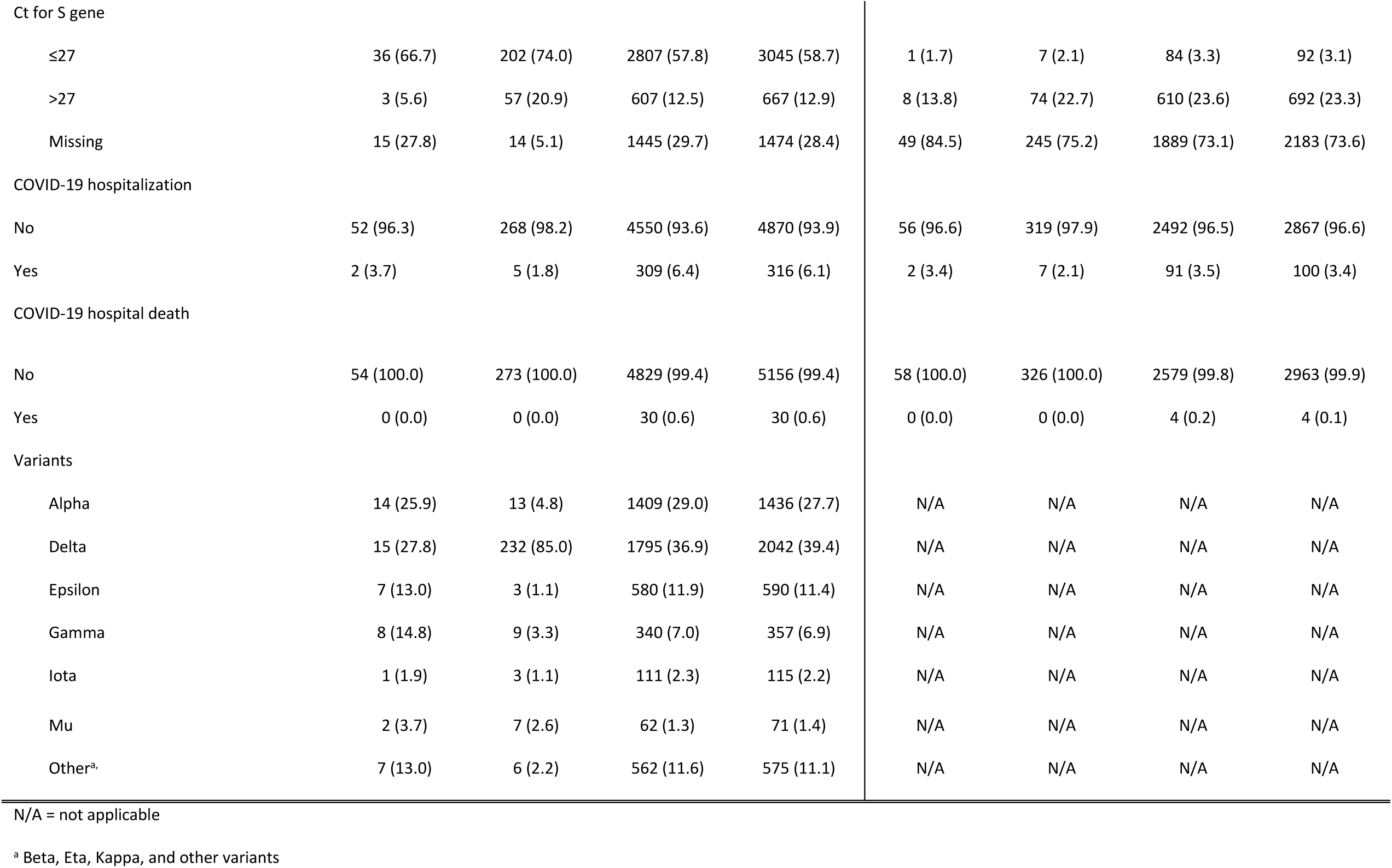
Sequencing characteristics of SARS-CoV-2 specimens, by sequencing status and mRNA-1273 vaccination.

|  | Sequencing Success |  |  |  | Sequencing Failure (Unidentified Variants) |  |  |  |
| --- | --- | --- | --- | --- | --- | --- | --- | --- |
|  | Vaccinated | Vaccinated | Unvaccinated | Total | Vaccinated | Vaccinated | Unvaccinated | Total |
|  | 1 dose | 2 dose |  |  | 1 dose | 2 dose |  |  |
|  | n (%) | n (%) | n (%) | n (%) | n (%) | n (%) | n (%) | n (%) |
| Overall, n | 54 | 273 | 4859 | 5186 | 58 | 326 | 2583 | 2967 |
| Specimen type |  |  |  |  |  |  |  |  |
| Nasopharyngeal/oropharyngeal swab | 53 (98.1) | 262 (96.0) | 4698 (96.7) | 5013 (96.7) | 48 (82.8) | 295 (90.5) | 2349 (90.9) | 2692 (90.7) |
| Saliva (asymptomatic) | 1 (1.9) | 11 (4.0) | 161 (3.3) | 173 (3.3) | 10 (17.2) | 31 (9.5) | 234 (9.1) | 275 (9.3) |
| Ct values for N, ORF1ab, and S genes |  |  |  |  |  |  |  |  |
| Any of 3 Ct values ≤27 | 48 (88.9) | 227 (83.2) | 4196 (86.4) | 4471 (86.2) | 3 (5.2) | 7 (2.1) | 148 (5.7) | 158 (5.3) |
| All non-missing Ct values >27 | 6 (11.1) | 46 (16.8) | 659 (13.6) | 711 (13.7) | 27 (46.6) | 167 (51.2) | 1780 (68.9) | 1974 (66.5) |
| All 3 Ct values missing | 0 (0.0) | 0 (0.0) | 4 (0.1) | 4 (0.1) | 28 (48.3) | 152 (46.6) | 655 (25.4) | 835 (28.1) |
| Ct for N gene |  |  |  |  |  |  |  |  |
| ≤27 | 45 (83.3) | 216 (79.1) | 4088 (84.1) | 4349 (83.9) | 3 (5.2) | 6 (1.8) | 132 (5.1) | 141 (4.8) |
| >27 | 9 (16.7) | 57 (20.9) | 765 (15.7) | 831 (16.0) | 21 (36.2) | 132 (40.5) | 1469 (56.9) | 1622 (54.7) |
| Missing | 0 (0.0) | 0 (0.0) | 6 (0.1) | 6 (0.1) | 34 (58.6) | 188 (57.7) | 982 (38.0) | 1204 (40.6) |
| Ct for ORF1ab gene |  |  |  |  |  |  |  |  |
| ≤27 | 45 (83.3) | 217 (79.5) | 4052 (83.4) | 4314 (83.2) | 1 (1.7) | 7 (2.1) | 108 (4.2) | 116 (3.9) |
| >27 | 9 (16.7) | 56 (20.5) | 800 (16.5) | 865 (16.7) | 28 (48.3) | 165 (50.6) | 1775 (68.7) | 1968 (66.3) |
| Missing | 0 (0.0) | 0 (0.0) | 7 (0.1) | 7 (0.1) | 29 (50.0) | 154 (47.2) | 700 (27.1) | 883 (29.8) |
| Ct for S gene |  |  |  |  |  |  |  |  |
| ≤27 | 36 (66.7) | 202 (74.0) | 2807 (57.8) | 3045 (58.7) | 1 (1.7) | 7 (2.1) | 84 (3.3) | 92 (3.1) |
| >27 | 3 (5.6) | 57 (20.9) | 607 (12.5) | 667 (12.9) | 8 (13.8) | 74 (22.7) | 610 (23.6) | 692 (23.3) |
| Missing | 15 (27.8) | 14 (5.1) | 1445 (29.7) | 1474 (28.4) | 49 (84.5) | 245 (75.2) | 1889 (73.1) | 2183 (73.6) |
| COVID-19 hospitalization |  |  |  |  |  |  |  |  |
| No | 52 (96.3) | 268 (98.2) | 4550 (93.6) | 4870 (93.9) | 56 (96.6) | 319 (97.9) | 2492 (96.5) | 2867 (96.6) |
| Yes | 2 (3.7) | 5 (1.8) | 309 (6.4) | 316 (6.1) | 2 (3.4) | 7 (2.1) | 91 (3.5) | 100 (3.4) |
| COVID-19 hospital death |  |  |  |  |  |  |  |  |
| No | 54 (100.0) | 273 (100.0) | 4829 (99.4) | 5156 (99.4) | 58 (100.0) | 326 (100.0) | 2579 (99.8) | 2963 (99.9) |
| Yes | 0 (0.0) | 0 (0.0) | 30 (0.6) | 30 (0.6) | 0 (0.0) | 0 (0.0) | 4 (0.2) | 4 (0.1) |
| Variants |  |  |  |  |  |  |  |  |
| Alpha | 14 (25.9) | 13 (4.8) | 1409 (29.0) | 1436 (27.7) | N/A | N/A | N/A | N/A |
| Delta | 15 (27.8) | 232 (85.0) | 1795 (36.9) | 2042 (39.4) | N/A | N/A | N/A | N/A |
| Epsilon | 7 (13.0) | 3 (1.1) | 580 (11.9) | 590 (11.4) | N/A | N/A | N/A | N/A |
| Gamma | 8 (14.8) | 9 (3.3) | 340 (7.0) | 357 (6.9) | N/A | N/A | N/A | N/A |
| Iota | 1 (1.9) | 3 (1.1) | 111 (2.3) | 115 (2.2) | N/A | N/A | N/A | N/A |
| Mu | 2 (3.7) | 7 (2.6) | 62 (1.3) | 71 (1.4) | N/A | N/A | N/A | N/A |
| Other<sup>a</sup> | 7 (13.0) | 6 (2.2) | 562 (11.6) | 575 (11.1) | N/A | N/A | N/A | N/A |

N/A = not applicable
<sup>a</sup> Beta, Eta, Kappa, and other variants

Among the 2,027 Delta cases matched to 10,135 controls (**Table 2**), 66.2% were aged 18-44 years, 55.9% were female, and 42.7% were Hispanic. Delta cases and controls had similar distributions of lung disease, autoimmune conditions, median neighborhood income, and KPSC physician/employee status. Compared to controls, Delta cases had lower comorbidity and frailty indices and less commonly had kidney disease, heart disease, liver disease, diabetes, immunocompromised status, pregnancy, and history of COVID-19 diagnosis, but more commonly had history of SARS-CoV-2 molecular testing and Medicaid. Delta cases also had fewer health care visits in the prior year than controls, and less commonly had preventive care. In addition, there were some differences by medical center, month of specimen collection, and specimen type. Characteristics of cases and matched controls for non-Delta and unidentified variants are described in **Tables S2-S8**.

**Table 2.**
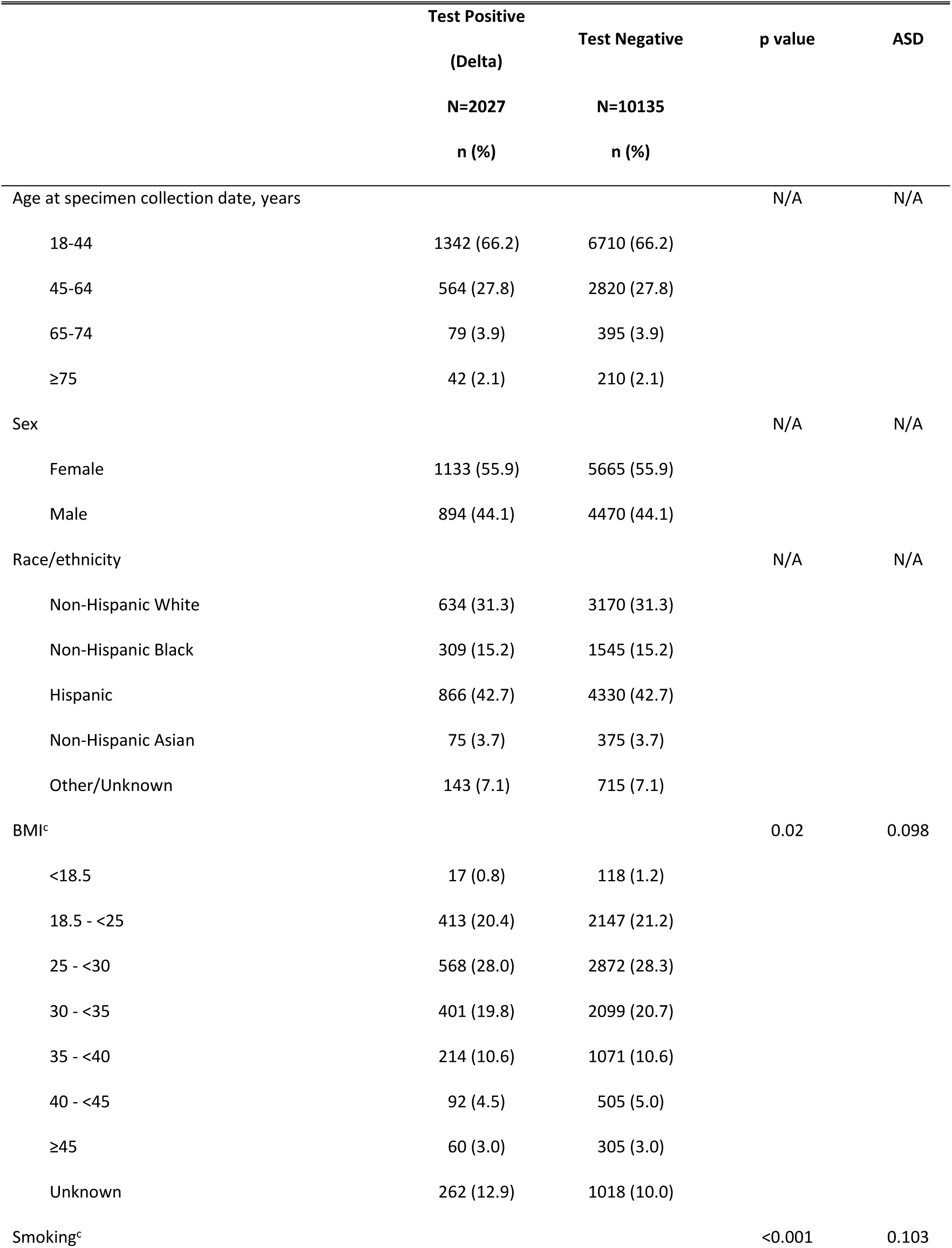
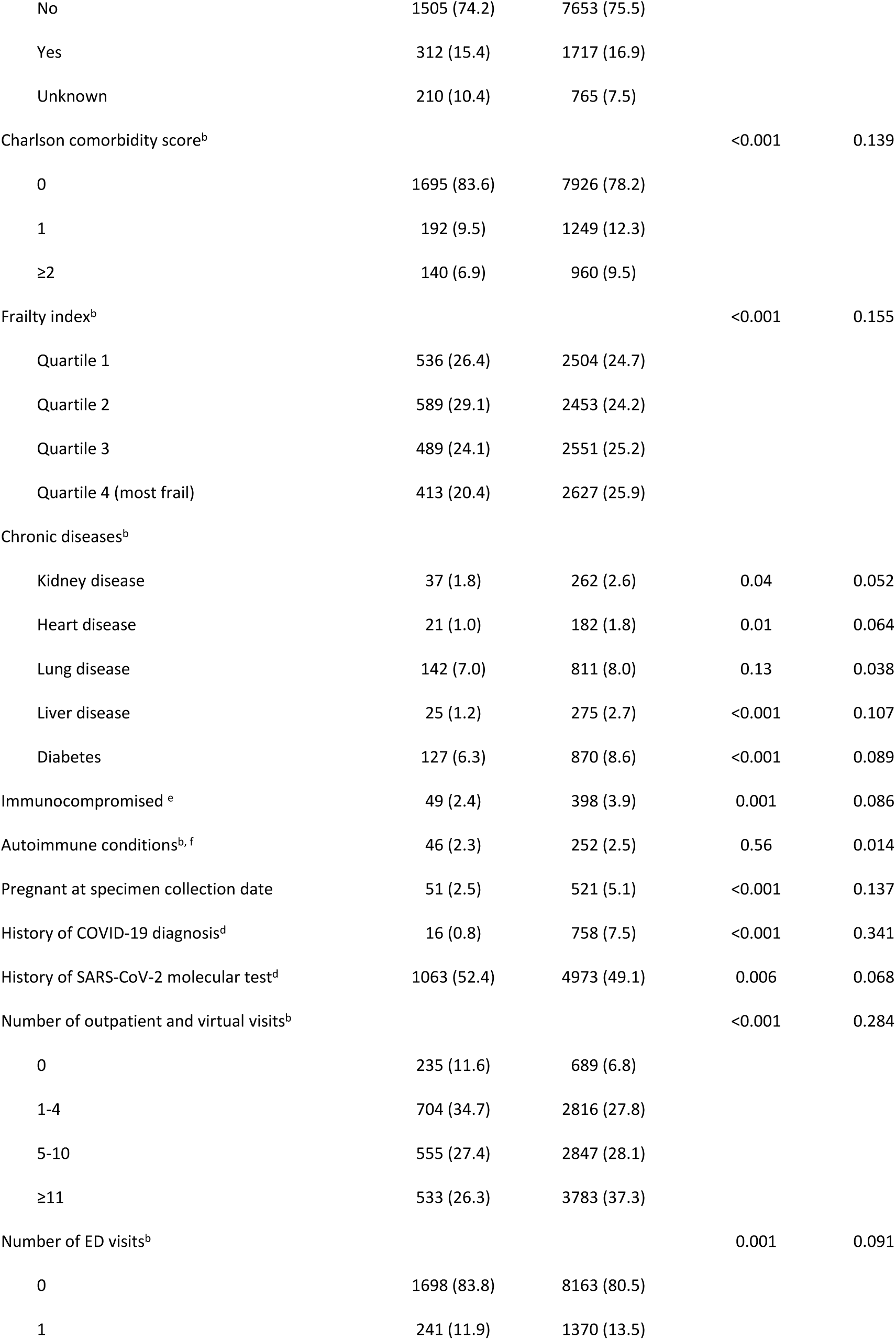
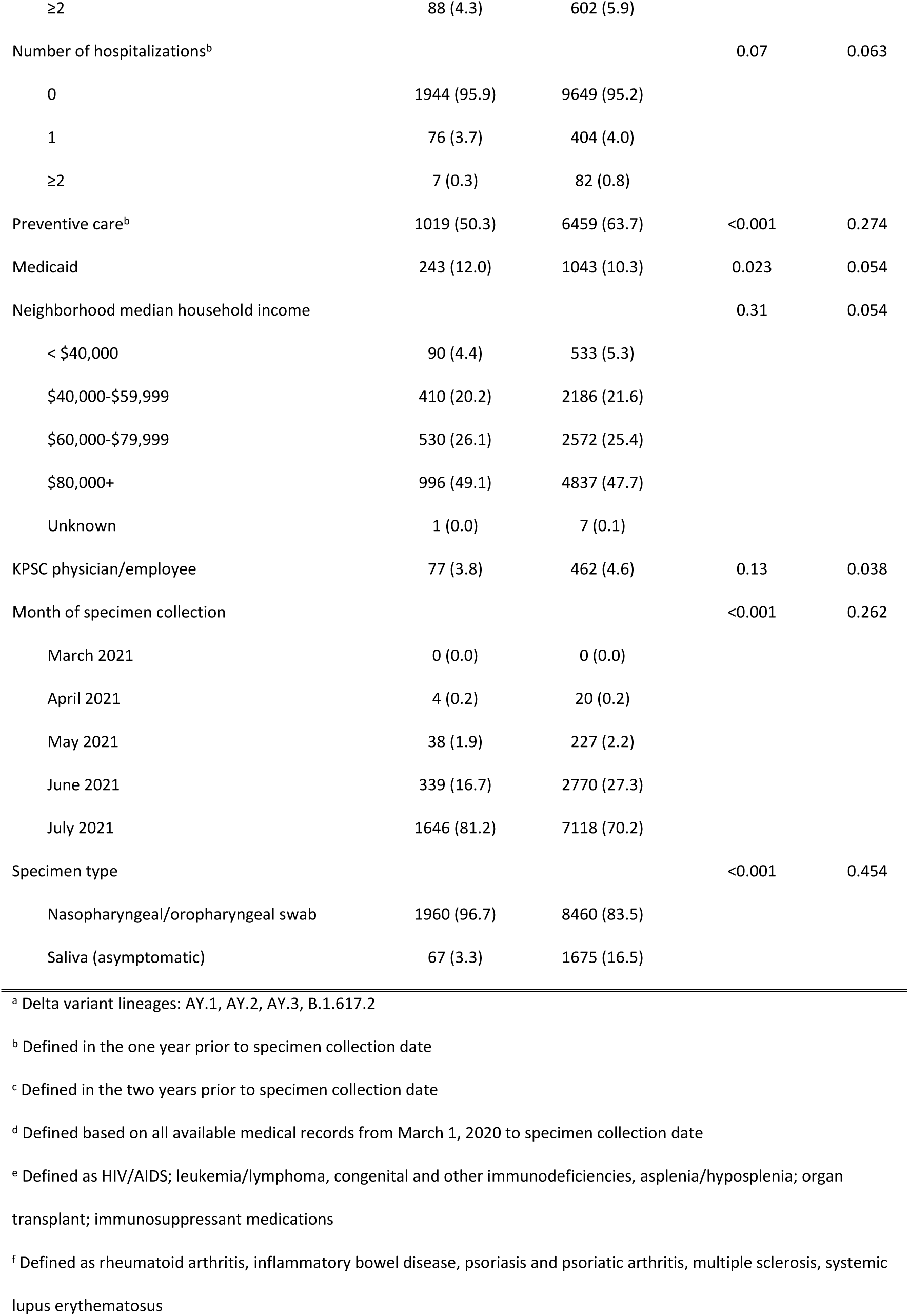
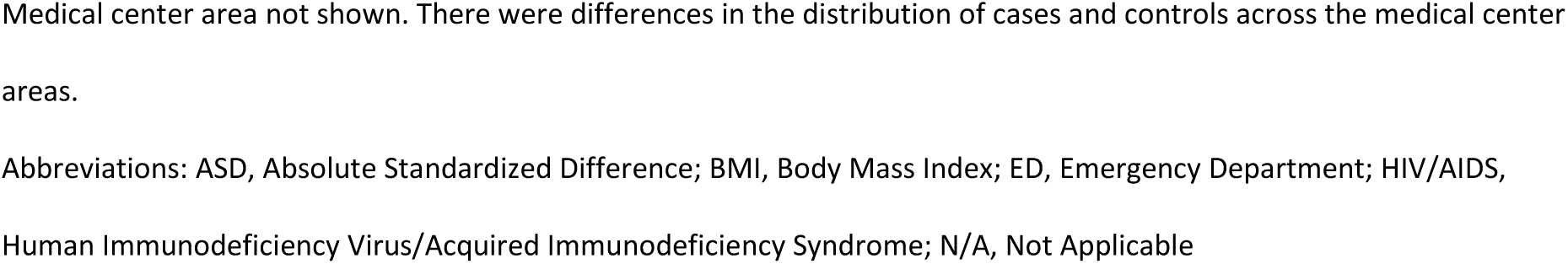
Characteristics of SARS-CoV-2 test-positive cases for Delta^a^ variant and test-negative controls (2-dose analysis)

Among Delta cases, 232 (11.4%) were fully vaccinated **(Figure 1 and Table S9)**. Among controls matched to Delta cases, 4,588 (45.3%) were fully vaccinated. In comparison, only 0.9% of Alpha cases and 24.4% of Alpha controls were fully vaccinated. VE (95% CI) against infection with Delta was 86.7% (84.3-88.7%), moderately lower than the high VE against Alpha (98.4% [96.9-99.1%]). VE against Mu was 90.4% (73.9-96.5%). VE against other identified non-Delta variants ranged from 95.5-97.6%, while VE against unidentified variants was 79.9% (76.9-82.5%). VE of 1 dose of mRNA-1273 was lower against all variants, ranging from 45.8% (0.0-88.9%) against Mu to 90.1% (82.9-94.2%) against Alpha (**Table S10**).

**Figure 1:**
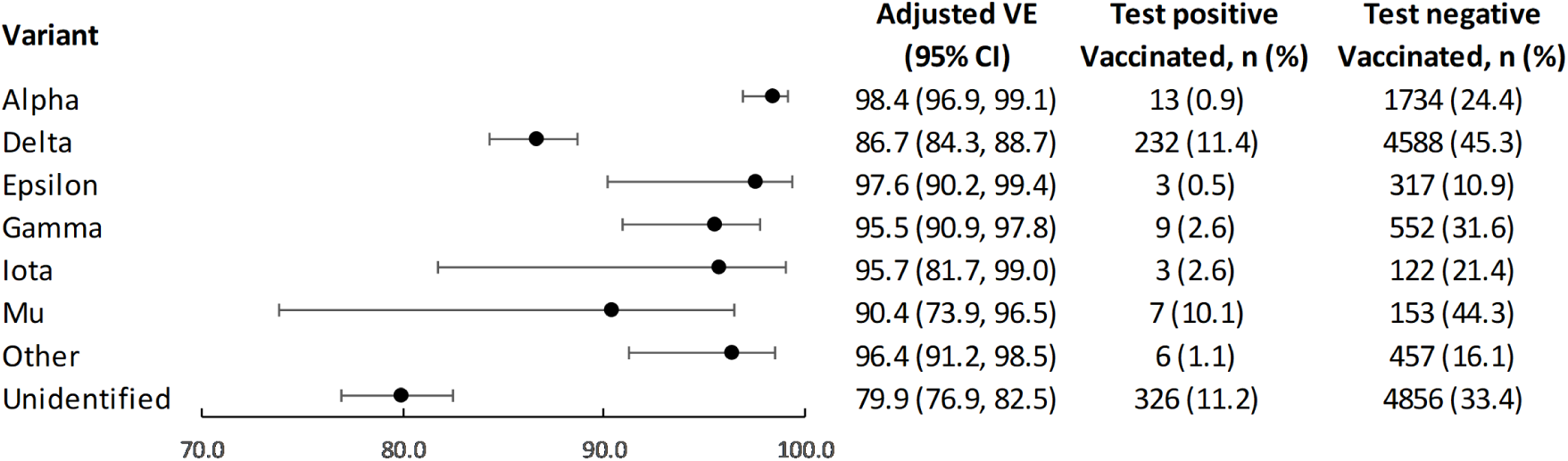
Vaccine effectiveness of 2 doses of mRNA-1273 against infection with SARS-CoV-2 variants.

In analyses of VE against Delta infection by time since receipt of second dose, VE was highest at 14-60 days (94.1% [90.5-96.3%]) and declined moderately, with VE of 80.0% (70.2-86.6%) at 151-180 days (**Figure 2 and Table S11**). VE against non-Delta infection also declined, with increasing time since vaccination, though not as sharply as for Delta (98.6% [97.3-99.3%] at 14-60 days to 88.7% [73.2-95.2%] at 121-150 days); analyses could not be done for later time points due to decline in prevalence. VE against unidentified variants was 83.6% (79.5-86.9%) at 14-60 days, declining to 68.5% (51.3-79.6%) at 151-180 days.

**Figure 2:**
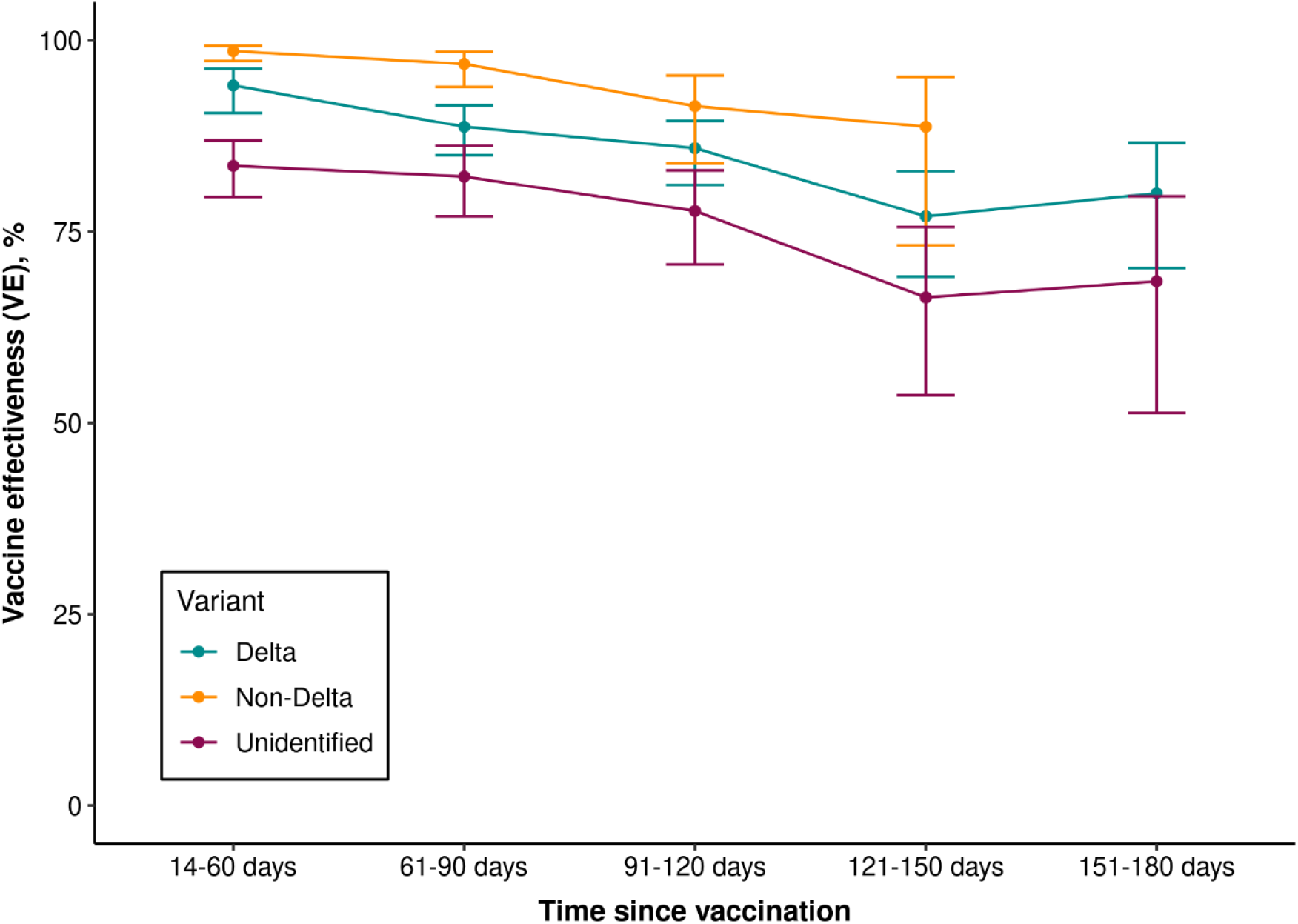
Vaccine effectiveness of 2 doses of mRNA-1273 against infection with SARS-CoV-2 variants by time since vaccination.

In analyses by age group, VE of 2 doses of mRNA-1273 against Delta infection was 87.9% (85.5-89.9%) among individuals aged 18-64 years and 75.2% (59.6-84.8%) among individuals aged ≥65 years (**Figure S3**). Among those aged 18-64 years, VE against Delta declined from 95.1% (91.8-97.1%) at 14-60 days to 79.4% (68.8-86.3%) at 151-180 days. Among individuals aged ≥65 years, confidence intervals were wide due to fewer cases in this age group, making trends less apparent.

However, during the study period robust protection was observed against hospitalization for Delta. Among hospitalized Delta cases and controls, 5 (3.5%) and 4,815 (40.1%) were fully vaccinated, respectively (**Table 3**). VE against hospitalization for Delta was high at 97.6% (92.8-99.2%). Similarly, VE against hospitalization for unidentified variants was 96.6% (89.4-98.9%). VE against hospitalization for non-Delta variants was not estimated due to 0 hospitalizations with non-Delta variants identified among vaccinated individuals.

**Table 3.** Vaccine effectiveness of 2 doses of mRNA-1273 against COVID-19 hospitalization with Delta, non-Delta or unidentified variants.

| Variant | Test Positive |  | Test Negative |  | Odds Ratio (95% CI) |  | Vaccine Effectiveness (95% CI) |  |
| --- | --- | --- | --- | --- | --- | --- | --- | --- |
|  | Vaccinated n (%) | Unvaccinated n (%) | Vaccinated n (%) | Unvaccinated n (%) | Unadjusted | Adjusted<sup>a</sup> | Unadjusted (%) | Adjusted<sup>a</sup> (%) |
| Delta | 5 (3.5) | 136 (96.5) | 4815 (40.1) | 7206 (59.9) | 0.03 (0.01, 0.07) | 0.02 (0.01, 0.07) | 97.4 (92.8, 99.0) | 97.6 (92.8, 99.2) |
| Non-Delta | 0 (0.0) | 173 (0.9) | 3376 (18.1) | 15081 (81.0) | N/E | N/E | N/E | N/E |
| Unidentified | 7 (7.1) | 91 (92.9) | 5175 (29.8) | 12181 (70.2) | 0.05 (0.02, 0.13) | 0.03 (0.01, 0.11) | 94.8 (86.8, 98.0) | 96.6 (89.4, 98.9) |

<sup>a</sup> Model adjustment:
Model for Delta variant adjusted for covariates: smoking, Charlson comorbidity score, frailty index, liver disease, pregnancy, history of COVID-19 diagnosis, number of outpatient and virtual visits, preventive care, medical center area, month of specimen collection, specimen type.
Model for Unidentified variants adjusted for covariates: BMI, smoking, Charlson comorbidity score, frailty index, pregnancy, history of COVID-19 diagnosis, number of outpatient and virtual visits, number of ED visits, preventive care, medical center area, month of specimen collection, specimen type.
Abbreviations: BMI, Body Mass Index; CI, Confidence Interval; ED, Emergency Department, N/E= Not Estimable

## DISCUSSION

This real-world study provides evidence of high VE of 2 doses of mRNA-1273 against multiple SARS-CoV-2 variants, including Delta. Although prior studies of BNT162b2 and other COVID-19 vaccines have examined VE against Delta,^13,29-33^ few studies of mRNA-1273 have reported variant-specific VE.^31,33^ Our study addresses this gap, finding that mRNA-1273 was protective against infection with Delta and other identified SARS-CoV-2 variants (Alpha, Epsilon, Gamma, Iota, Mu, and others). VE against Delta infection was moderately lower than VE against identified non-Delta variants (86.7% vs. 90.4-98.4%), as observed in several other studies.^13,30^

During the Delta phase of the pandemic, breakthrough infections among fully vaccinated individuals have occurred,^16 17^ but VE of COVID-19 vaccines against severe disease has remained robust.^18,19,31,34^ In our study, VE against hospitalization with Delta was high (97.6%). Only 5 fully vaccinated Delta cases were hospitalized, no fully vaccinated non-Delta cases were hospitalized, and no hospitalized deaths occurred among any fully vaccinated cases. This finding is consistent with prior studies suggesting that fully vaccinated individuals with breakthrough infections tend to have attenuated viral load, fewer symptoms, and shorter illness duration, though severe outcomes can still occur.^7,35,36^

In this study, variants were unidentified for the substantial proportion of specimens that failed sequencing, possibly due to lower viral loads, timing of specimen collection relative to symptom onset, or poor specimen quality. The higher proportion of failed vs. successfully sequenced specimens that were fully vaccinated, saliva specimens (used only for asymptomatic testing at KPSC), and specimens with higher Ct values, altogether suggest less severe disease, which may help explain the lower VE against infection observed for this group (79.9%). Despite the lower VE against infection, VE against hospitalization with unidentified variants remained high (96.6%).

Our study identified modest waning of mRNA-1273 VE against Delta infection; VE decreased from 94.1% in the 14-60 days after vaccination to 80.0% in the 151-180 days after vaccination. Similar reductions were observed among individuals aged 18-64 years, but among those aged ≥65 years, 95% CIs for VE by time since vaccination were wide. Other observational studies have found reduced VE of mRNA-based vaccines against infection in periods before and after Delta became predominant, some of which identified steeper declines than observed in our study.^20,21,29,32^ We also identified a decline in VE of mRNA-1273 against non-Delta infections but this reduction was less pronounced than for Delta or for unidentified variants. Declines in VE might also be partly due to differences in characteristics and behaviors of individuals vaccinated earlier vs. later in phased vaccine rollout.

Our real-world findings complement existing immunogenicity and Phase 3 trial follow-up data of mRNA-1273 protection against SARS-CoV-2 variants, including Delta. High levels of neutralizing antibodies to Delta and other variants were elicited following 2-doses of mRNA-1273.^37^ These antibodies were found to persist 6 months after vaccination, albeit at reduced levels compared with peak activity. Among Phase 3 trial participants, incidence rates of COVID-19 and severe COVID-19 during the months when Delta was predominant were lower among those who were vaccinated with mRNA-1273 more recently (median 8 months after first dose) compared to those vaccinated initially (median 13 months after first dose).^38^

The findings of this study have implications for booster doses, which have been authorized in certain populations. Questions remain over the benefits of booster doses in different populations and optimal approaches for boosting immunity. Several studies have identified higher VE for mRNA-1273 after 1 and 2 doses compared to BNT162b2,^12,39^ suggesting different booster strategies could be appropriate depending on product. However, efforts to deploy booster doses must not replace efforts to reach unvaccinated individuals, who comprise most COVID-19 hospitalizations and deaths. Booster dose strategies must also prioritize global vaccine production and allocation.

Our study had multiple strengths. We systematically collected positive SARS-CoV-2 specimens across KPSC care settings and sent specimens for WGS regardless of Ct value. Test-positive cases were matched to test-negative controls on demographic factors and calendar time, reducing secular confounding due to differences over time in transmission, vaccination rollout, and testing. We examined mRNA-1273 VE against multiple variants, including Delta and Mu. We also evaluated VE against Delta hospitalization. With data up to 6 months following receipt of two doses of mRNA-1273, we stratified analyses of duration of protection by variant type and age group.

Our study also had several limitations. Although test-negative designs might reduce bias due to factors associated with care-seeking,^40^ this design was generalizable to individuals who were tested and was therefore less generalizable to individuals with mild or no symptoms who did not seek testing. The detailed KPSC EHR enabled adjustment for comprehensive sociodemographic and clinical covariates, but there could still be residual confounding due to unmeasured factors associated with both testing and vaccination. Misclassification of case/control status could occur due to false positives or negatives, although sensitivity and specificity of PCR testing was high. Misclassification of vaccine exposure was also possible but unlikely due to comprehensive KPSC and external COVID-19 vaccination records. Sample size was limited in the subgroup aged ≥65 years for the analysis of VE against Delta infection by time since vaccination.

In conclusion, this study found high VE of mRNA-1273 against infection due to SARS-CoV-2 variants, including Delta, adding to the limited literature specific for mRNA-1273. VE against hospitalization for Delta was also high. This study provides reassuring evidence of the effectiveness of 2 doses of mRNA-1273 in preventing infection and COVID-19 hospitalization due to variants including Delta. Moderate declines in VE were observed against Delta infection. Additional research is required to inform booster dose strategies over time.

## Data Availability

Individual-level data reported in this study are not publicly shared. Upon request, and subject to review, KPSC may provide the deidentified aggregate-level data that support the findings of this study. Deidentified data (including participant data as applicable) may be shared upon approval of an analysis proposal and a signed data access agreement.

## AUTHOR CONTRIBUTIONS

Conceptualization, KJB, LSS, LQ, CAT, HFT; acquisition, analysis, or interpretation of data, KJB, LSS, LQ, BKA, YL, AF, JHK, CAT, HFT; Drafting of the manuscript, KJB, LSS; Critical revision of the manuscript for important intellectual content, LQ, BKA, YL, GSL, YT, AF, MA, JET, HST, JHK, YDP, CAT, HFT; Statistical analysis, LQ, YL, YT, JET; Obtained funding, CAT, HFT; Administrative, technical, or material support, LSS, GSL, MA, HST, CAT, YDP; Supervision, CAT, HFT. All authors approved the final version of the manuscript.

## REQUIRED STATEMENTS

1. Research protocol was approved by KPSC IRB
2. Waiver of informed consent obtained from KPSC IRB due to research posing no more than minimal risk to subjects
3. Registration number and registry name in text – Not applicable
4. Accession numbers and repository names for studies containing microarrays – Not applicable
5. Name of person who analyzed the data – Lei Qian and Yi Luo

## ACKNOWLEDGMENTS

This study was funded by Moderna Inc. Medical writing and editorial assistance was provided by Srividya Ramachandran, PhD, and Jared Mackenzie, PhD, of MEDiSTRAVA in accordance with Good Publication Practice (GPP3) guidelines, funded by Moderna Inc, and under the direction of the authors. The authors would like to acknowledge the following Kaiser Permanente Southern California staff: Donald Kaplan, PharmD, Daniel Ehrlich, PharmD, Dale Timothy, PharmD, Patrick Kerrigan, PharmD, David Cheng, PharmD, Kevin Ohara, Pharm D, Joel Christian, PharmD, Danny Byun, PharmD, Erin Matsushita, PharmD, Dennis Curtis, PharmD, Victoria Hong, PharmD, and Arthur Librea, PharmD, for their role in COVID-19 vaccine logistics and coordination; Soon Kyu Choi, Jennifer Charter, Joy Gelfond, Radha Bathala, and Lee Childs for their coordination in processing SARS-CoV-2 specimens; and Raul Calderon, Kourtney Kottman, Ana Acevedo, Elmer Ayala, and Jonathan Arguello for their technical and laboratory support in processing SARS-CoV-2 specimens. The authors would like to acknowledge Helix OpCo, LLC, for their whole genome sequencing of SARS-CoV-2 specimens. The authors would also like to acknowledge the contributions by Moderna staff: Groves Dixon, PhD, and Julie Vanas. The authors thank the patients of Kaiser Permanente for their partnership with us to improve their health. Their information, collected through our electronic health record systems, leads to findings that help us improve care for our members and can be shared with the larger community.

