## Supplementary for "Effectiveness of mRNA-1273 against Delta, Mu, and other emerging variants"

**SUPPLEMENTARY APPENDIX**

**Authors:** Katia J. Bruxvoort, PhD,[1,2] Lina S. Sy, MPH,[1] Lei Qian, PhD,[1] Bradley K. Ackerson, MD,[1] Yi Luo, PhD,[1] Gina S. Lee, MPH,[1] Yun Tian, MS,[1] Ana Florea, PhD,[1] Michael Aragones, MD,[1] Julia E. Tubert, MPH,[1] Harpreet S. Takhar, MPH,[1] Jennifer Ku, PhD,[1] Yamuna D. Paila, PhD,[3] Carla A. Talarico, PhD,[3] Hung Fu Tseng, PhD[1,4]

**Affiliations:** [1]Kaiser Permanente Southern California, 100 S. Los Robles Ave., Pasadena, CA 91101, USA; [2]University of Alabama at Birmingham, 1665 University Blvd, Birmingham, AL 35233, USA; [3]Moderna Inc., 200 Technology Square, Cambridge, MA 02139, USA; [4]Kaiser Permanente Bernard J. Tyson School of Medicine, 98 S. Los Robles Ave., Pasadena, CA 91101, USA

### Supplementary Methods

**Whole genome sequencing approach**

KPSC began sending all positive SARS-CoV-2 specimens to a commercial laboratory (Helix, California, USA) for whole genome sequencing (WGS) beginning March 2021. In Helix workflow, SARS-CoV-2 genome capture was accomplished using hybridization capture with the xGen COVID-19 Capture Panel (Integrated DNA Technologies, Iowa, USA). Specimens were sequenced using the NovaSeq 6000 Sequencing System S1 Reagent Kit v1.5 (300 cycles) (Ilumina, California, USA). Sequence reads were aligned to the SARS-CoV-2 reference genome (NCBI accession NC_045512.2) and the human transcriptome (GENCODE v37) using BWA-MEM, and variants were called using the Haplotyper algorithm (Sentieon Inc, California, USA). Consensus sequences (FASTA) required each allele to have at least 5 unique reads with at least 80% of the reads supporting the allele.

**Power calculations**

Analyses were conducted for variants with at least 20 cases, based on the following power calculations. For a 1:5 matched test-negative design, assuming a 60% vaccination rate (2 doses of mRNA-1273) in test-negative controls, 20 cases of were needed to detect a vaccine effectiveness of 0.8 with 80% statistical power.

Table S1. ICD-10 diagnosis codes for subgroup conditions and for covariates to be considered as potential confounders

| Covariate | ICD-10 codes |
|---|---|
| **Immunosuppressant conditions** | |
| Lymphoma/leukemia | C81.*C86.*, C88.*, C90.*-C96.*, D45, D46.* |
| Congenital and other immunodeficiencies | D61.09, D61.3, D61.82, D61.9, D70.0, D71, D80.0, D80.1, D80.5, D80.8, D81.*, D82.*, D83.*, D84.0, D84.1, D84.89, D84.9, D89.81*, D89.82, D89.9, E31.0, E70.330, G11.3, Q82.4, Q89.0* |
| Asplenia/hyposplenia | D57.00, D57.01, D57.02, D57.1, D57.2, D57.20, D57.21, D57.211, D57.212, D57.219, D57.4, D57.40, D57.41, D57.411, D57.412, D57.419, D57.8, D57.80, D57.81, D57.811, D57.812, D57.819, D73.0, Q89.01, Q89.09, Z90.81 |
| **Autoimmune conditions** | |
| Rheumatic disease | M05.*, M06.*, M31.5, M32.*-M34.*, M35.1, M35.3, M36.0 |
| Inflammatory bowel disease | K50.*, K51.* |
| Psoriasis | L40.* |
| Psoriatic arthritis | L40.5* |
| Multiple sclerosis | G35 |
| Systemic lupus erythematosus | M32.1, M32.8, M32.9 |
| **Charlson comorbidity** | |
| Myocardial Infarction | I21.*, I22.*, I25.2 |
| Congestive Heart Failure | I09.9, I11.0, I13.0, I13.2, I25.5, I42.0, I42.5-I42.9, I43.*, I50.*, P29.0 |
| Peripheral Vascular Disease | I70.*, I71.*, I73.1, I73.8*, I73.9, I77.1, I79.0, I79.2, K55.1, K55.8, K55.9, Z95.8*, Z95.9 |
| Cerebrovascular Disease | G45.*, G46.*, H34.0*, I60.*-I69.* |
| Connective tissue disease | M05.*, M06.*, M31.5, M32.*-M34.*, M35.1, M35.3, M36.0 |
| Dementia | F00.*-F03.*, F05.1, G30.*, G31.1 |
| Chronic pulmonary disease | I27.8*, I27.9, J40.*-J47.*, J60.*-J67.*, J68.4, J70.1, J70.3 |
| Peptic ulcer disease | K25.*-K28.* |
| Diabetes mellitus | E10.*-E14.* |
| Paraplegia and hemiplegia | G04.1, G11.4, G80.1, G80.2, G81.*, G82.*, G83.0, G83.1*-G83.3*, G83.4, G83.9 |
| Renal disease | I12.0, I12.9, I13.1*, N03.2-N03.7, N05.2-N05.7, N18.*, N19.*, N25.0, Z49.0*-Z49.2*, Z94.0, Z99.2 |
| Liver disease | B18.*, I85.0*, I86.4, I98.2, K70.0, K70.1*-K70.4*, K70.9, K71.1*, K71.3, K71.4, K71.5*, K71.7, K72.1*, K72.9*, K73.*, K74.*, K76.0, K76.2-K76.7, K76.8*, K76.9, Z94.4 |
| Cancer: any malignancy, including lymphoma and leukemia, except malignant neoplasm of skin | C00.*-C26.*, C30.*-C34.*, C37.*-C41.*, C43.*, C45.*-C58.*, C60.*-C76.*, C81.*-C85.*, C88.*, C90.*-C97.* |
| Metastatic Solid Tumor | C77.*-C80.* |
| **Chronic diseases** | |
| Kidney disease | I12.0, I12.9, I13.1*, N03.2-N03.7, N05.2-N05.7, N18.*, N19.*, N25.0, Z49.0*-Z49.2*, Z94.0, Z99.2 |
| Heart disease | I21.*, I22.*, I25.2, I09.9, I11.0, I13.0, I13.2, I25.5, I42.0, I42.5-I42.9, I43.*, I50.*, P29.0 |
| Lung disease | I27.8*, I27.9, J40.*-J47.*, J60.*-J67.*, J68.4, J70.1, J70.3 |
| Liver disease | B18.*, I85.0*, I86.4, I98.2, K70.0, K70.1*-K70.4*, K70.9, K71.1*, K71.3, K71.4, K71.5*, K71.7, K72.1*, K72.9*, K73.*, K74.*, K76.0, K76.2-K76.7, K76.8*, K76.9, Z94.4 |
| Diabetes mellitus | E10.*-E14.* |

Table S2. Characteristics of SARS-CoV-2 test-positive Alpha[a] cases and test-negative controls (2-dose analysis)

| | Test Positive (Alpha) N=1422 n (%) | Test Negative N=7110 n (%) | p value | ASD |
|---|---|---|---|---|
| Age at specimen collection date, years | | | N/A | N/A |
| 18-44 | 839 (59.0) | 4195 (59.0) | | |
| 45-64 | 498 (35.0) | 2490 (35.0) | | |
| 65-74 | 64 (4.5) | 320 (4.5) | | |
| ≥75 | 21 (1.5) | 105 (1.5) | | |
| Sex | | | N/A | N/A |
| Female | 780 (54.9) | 3900 (54.9) | | |
| Male | 642 (45.1) | 3210 (45.1) | | |
| Race/ethnicity | | | N/A | N/A |
| Non-Hispanic White | 479 (33.7) | 2395 (33.7) | | |
| Non-Hispanic Black | 189 (13.3) | 945 (13.3) | | |
| Hispanic | 588 (41.4) | 2940 (41.4) | | |
| Non-Hispanic Asian | 56 (3.9) | 280 (3.9) | | |
| Other/Unknown | 110 (7.7) | 550 (7.7) | | |
| BMI[c] | | | <0.0001 | 0.1651 |
| <18.5 | 8 (0.6) | 94 (1.3) | | |
| 18.5 - <25 | 269 (18.9) | 1472 (20.7) | | |
| 25 - <30 | 361 (25.4) | 2005 (28.2) | | |
| 30 - <35 | 300 (21.1) | 1487 (20.9) | | |
| 35 - <40 | 165 (11.6) | 792 (11.1) | | |
| 40 - <45 | 70 (4.9) | 361 (5.1) | | |
| ≥45 | 61 (4.3) | 242 (3.4) | | |
| Unknown | 188 (13.2) | 657 (9.2) | | |
| Smoking[c] | | | 0.0003 | 0.1153 |
| No | 1040 (73.1) | 5185 (72.9) | | |
| Yes | 232 (16.3) | 1371 (19.3) | | |
| Unknown | 150 (10.5) | 554 (7.8) | | |
| Charlson comorbidity score[b] | | | <0.0001 | 0.1612 |
| 0 | 1152 (81.0) | 5355 (75.3) | | |
| 1 | 165 (11.6) | 916 (12.9) | | |
| ≥2 | 105 (7.4) | 839 (11.8) | | |
| Frailty index[b] | | | <0.0001 | 0.1923 |
| Quartile 1 | 306 (21.5) | 1693 (23.8) | | |
| Quartile 2 | 465 (32.7) | 1802 (25.3) | | |
| Quartile 3 | 366 (25.7) | 1767 (24.9) | | |
| Quartile 4 (most frail) | 285 (20.0) | 1848 (26.0) | | |
| Chronic diseases[b] | | | | |
| Kidney disease | 29 (2.0) | 250 (3.5) | 0.0043 | 0.0900 |
| Heart disease | 20 (1.4) | 181 (2.5) | 0.0097 | 0.0819 |
| Lung disease | 89 (6.3) | 644 (9.1) | 0.0006 | 0.1054 |
| Liver disease | 26 (1.8) | 231 (3.2) | 0.0042 | 0.0904 |
| Diabetes | 132 (9.3) | 700 (9.8) | 0.5139 | 0.0191 |
| Immunocompromised[e] | 28 (2.0) | 305 (4.3) | <0.0001 | 0.1336 |
| Autoimmune conditions[b,f] | 29 (2.0) | 191 (2.7) | 0.1600 | 0.0426 |
| Pregnant at specimen collection date | 28 (2.0) | 384 (5.4) | <0.0001 | 0.1829 |
| History of COVID-19 diagnosis[d] | 6 (0.4) | 538 (7.6) | <0.0001 | 0.3711 |
| History of SARS-CoV-2 molecular test[d] | 727 (51.1) | 3644 (51.3) | 0.9305 | 0.0025 |
| Number of outpatient and virtual visits[b] | | | <0.0001 | 0.3458 |
| 0 | 182 (12.8) | 545 (7.7) | | |
| 1-4 | 511 (35.9) | 1863 (26.2) | | |
| 5-10 | 358 (25.2) | 1848 (26.0) | | |
| ≥11 | 371 (26.1) | 2854 (40.1) | | |
| Number of ED visits[b] | | | <0.0001 | 0.1851 |
| 0 | 1219 (85.7) | 5620 (79.0) | | |
| 1 | 148 (10.4) | 990 (13.9) | | |
| ≥2 | 55 (3.9) | 500 (7.0) | | |
| Number of hospitalizations[b] | | | 0.0007 | 0.1233 |

| | | | | |
|---|---|---|---|---|
| 0 | 1368 (96.2) | 6670 (93.8) | | |
| 1 | 47 (3.3) | 337 (4.7) | | |
| ≥2 | 7 (0.5) | 103 (1.4) | | |
| Preventive care[b] | 686 (48.2) | 4543 (63.9) | <0.0001 | 0.3194 |
| Medicaid | 164 (11.5) | 764 (10.7) | 0.3838 | 0.0250 |
| Neighborhood median household income | | | 0.1718 | 0.0729 |
| < $40,000 | 73 (5.1) | 397 (5.6) | | |
| $40,000-$59,999 | 278 (19.5) | 1517 (21.3) | | |
| $60,000-$79,999 | 399 (28.1) | 1790 (25.2) | | |
| $80,000+ | 670 (47.1) | 3399 (47.8) | | |
| Unknown | 2 (0.1) | 7 (0.1) | | |
| KPSC physician/employee | 47 (3.3) | 299 (4.2) | 0.1162 | 0.0474 |
| Month of specimen collection | | | <0.0001 | 0.1477 |
| March 2021 | 345 (24.3) | 1857 (26.1) | | |
| April 2021 | 532 (37.4) | 2269 (31.9) | | |
| May 2021 | 345 (24.3) | 1950 (27.4) | | |
| June 2021 | 126 (8.9) | 762 (10.7) | | |
| July 2021 | 74 (5.2) | 272 (3.8) | | |
| Specimen type | | | <0.0001 | 0.4551 |
| NP/OP swab | 1376 (96.8) | 5940 (83.5) | | |
| Saliva (asymptomatic) | 46 (3.2) | 1170 (16.5) | | |

[a] Alpha variant lineage: B.1.1.7
[b] Defined in the one year prior to specimen collection date
[c] Defined in the two years prior to specimen collection date
[d] Defined based on all available medical records from March 1, 2020 to specimen collection date
[e] Defined as HIV/AIDS; leukemia/lymphoma, congenital and other immunodeficiencies, asplenia/hyposplenia; organ transplant; immunosuppressant medications
[f] Defined as rheumatoid arthritis, inflammatory bowel disease, psoriasis and psoriatic arthritis, multiple sclerosis, systemic lupus erythematosus
Medical center area not shown. There were differences in the distribution of cases and controls across the medical center areas.
Abbreviations: ASD, Absolute Standardized Difference; BMI, Body Mass Index; ED, Emergency Department; HIV/AIDS, Human Immunodeficiency Virus/Acquired Immunodeficiency Syndrome; NP/OP, nasopharyngeal/oropharyngeal

Table S3. Characteristics of SARS-CoV-2 test-positive Epsilon[a] cases and test-negative controls (2-dose analysis)

| | Test Positive (Epsilon) N=583 n (%) | Test Negative N=2915 n (%) | p value | ASD |
|---|---|---|---|---|
| Age at specimen collection date, years | | | N/A | N/A |
| 18-44 | 346 (59.3) | 1730 (59.3) | | |
| 45-64 | 216 (37.0) | 1080 (37.0) | | |
| 65-74 | 16 (2.7) | 80 (2.7) | | |
| ≥75 | 5 (0.9) | 25 (0.9) | | |
| Sex | | | N/A | N/A |
| Female | 303 (52.0) | 1515 (52.0) | | |
| Male | 280 (48.0) | 1400 (48.0) | | |
| Race/ethnicity | | | N/A | N/A |
| Non-Hispanic White | 174 (29.8) | 870 (29.8) | | |
| Non-Hispanic Black | 34 (5.8) | 170 (5.8) | | |
| Hispanic | 307 (52.7) | 1535 (52.7) | | |
| Non-Hispanic Asian | 35 (6.0) | 175 (6.0) | | |
| Other/Unknown | 33 (5.7) | 165 (5.7) | | |
| BMI[c] | | | <0.0001 | 0.2537 |
| <18.5 | 6 (1.0) | 36 (1.2) | | |
| 18.5 - <25 | 105 (18.0) | 616 (21.1) | | |
| 25 - <30 | 161 (27.6) | 882 (30.3) | | |
| 30 - <35 | 117 (20.1) | 625 (21.4) | | |
| 35 - <40 | 76 (13.0) | 326 (11.2) | | |
| 40 - <45 | 21 (3.6) | 129 (4.4) | | |
| ≥45 | 14 (2.4) | 94 (3.2) | | |
| Unknown | 83 (14.2) | 207 (7.1) | | |
| Smoking[c] | | | <0.0001 | 0.2275 |
| No | 427 (73.2) | 2151 (73.8) | | |
| Yes | 88 (15.1) | 589 (20.2) | | |
| Unknown | 68 (11.7) | 175 (6.0) | | |
| Charlson comorbidity score[b] | | | <0.0001 | 0.2896 |
| 0 | 497 (85.2) | 2152 (73.8) | | |
| 1 | 50 (8.6) | 403 (13.8) | | |
| ≥2 | 36 (6.2) | 360 (12.3) | | |
| Frailty index[b] | | | <0.0001 | 0.3165 |
| Quartile 1 | 149 (25.6) | 726 (24.9) | | |
| Quartile 2 | 204 (35.0) | 666 (22.8) | | |
| Quartile 3 | 131 (22.5) | 748 (25.7) | | |
| Quartile 4 (most frail) | 99 (17.0) | 775 (26.6) | | |
| Chronic diseases[b] | | | | |
| Kidney disease | 8 (1.4) | 96 (3.3) | 0.0127 | 0.1275 |
| Heart disease | 3 (0.5) | 75 (2.6) | 0.0021 | 0.1675 |
| Lung disease | 27 (4.6) | 268 (9.2) | 0.0003 | 0.1806 |
| Liver disease | 13 (2.2) | 111 (3.8) | 0.0600 | 0.0923 |
| Diabetes | 40 (6.9) | 307 (10.5) | 0.0068 | 0.1305 |
| Immunocompromised[e] | 14 (2.4) | 137 (4.7) | 0.0127 | 0.1244 |
| Autoimmune conditions[b,f] | 20 (3.4) | 86 (3.0) | 0.5369 | 0.0273 |
| Pregnant at specimen collection date | 11 (1.9) | 140 (4.8) | 0.0016 | 0.1627 |
| History of COVID-19 diagnosis[d] | 2 (0.3) | 220 (7.5) | <0.0001 | 0.3766 |
| History of SARS-CoV-2 molecular test[d] | 299 (51.3) | 1678 (57.6) | 0.0052 | 0.1263 |
| Number of outpatient and virtual visits[b] | | | <0.0001 | 0.4151 |
| 0 | 65 (11.1) | 164 (5.6) | | |
| 1-4 | 225 (38.6) | 783 (26.9) | | |
| 5-10 | 152 (26.1) | 773 (26.5) | | |
| ≥11 | 141 (24.2) | 1195 (41.0) | | |
| Number of ED visits[b] | | | <0.0001 | 0.3108 |
| 0 | 508 (87.1) | 2243 (76.9) | | |

| | | | | |
|---|---|---|---|---|
| 1 | 61 (10.5) | 424 (14.5) | | |
| ≥2 | 14 (2.4) | 248 (8.5) | | |
| Number of hospitalizations[b] | | | 0.0151 | 0.1496 |
| 0 | 556 (95.4) | 2693 (92.4) | | |
| 1 | 24 (4.1) | 166 (5.7) | | |
| ≥2 | 3 (0.5) | 56 (1.9) | | |
| Preventive care[b], n() | 314 (53.9) | 1920 (65.9) | <0.0001 | 0.2468 |
| Medicaid | 59 (10.1) | 284 (9.7) | 0.7797 | 0.0126 |
| Neighborhood median household income | | | 0.8642 | 0.0572 |
| < $40,000 | 25 (4.3) | 149 (5.1) | | |
| $40,000-$59,999 | 122 (20.9) | 611 (21.0) | | |
| $60,000-$79,999 | 162 (27.8) | 778 (26.7) | | |
| $80,000+ | 274 (47.0) | 1375 (47.2) | | |
| Unknown | 0 (0.0) | 2 (0.1) | | |
| KPSC physician/employee | 16 (2.7) | 129 (4.4) | 0.0631 | 0.0905 |
| Month of specimen collection | | | 0.2761 | 0.0977 |
| March 2021 | 463 (79.4) | 2310 (79.2) | | |
| April 2021 | 110 (18.9) | 516 (17.7) | | |
| May 2021 | 8 (1.4) | 79 (2.7) | | |
| June 2021 | 2 (0.3) | 10 (0.3) | | |
| July 2021 | 0 (0.0) | 0 (0.0) | | |
| Specimen type | | | <0.0001 | 0.4285 |
| NP/OP swab | 556 (95.4) | 2394 (82.1) | | |
| Saliva (asymptomatic) | 27 (4.6) | 521 (17.9) | | |

[a] Epsilon variant lineages: B.1.427, B.1.429
[b] Defined in the one year prior to specimen collection date
[c] Defined in the two years prior to specimen collection date
[d] Defined based on all available medical records from March 1, 2020 to specimen collection date
[d] Defined as HIV/AIDS; leukemia/lymphoma, congenital and other immunodeficiencies, asplenia/hyposplenia; organ transplant; immunosuppressant medications
[f] Defined as rheumatoid arthritis, inflammatory bowel disease, psoriasis and psoriatic arthritis, multiple sclerosis, systemic lupus erythematosus
Medical center area not shown. There were differences in the distribution of cases and controls across the medical center areas.
Abbreviations: ASD, Absolute Standardized Difference; BMI, Body Mass Index; ED, Emergency Department; HIV/AIDS, Human Immunodeficiency Virus/Acquired Immunodeficiency Syndrome; NP/OP, nasopharyngeal/oropharyngeal

Table S4. Characteristics of SARS-CoV-2 test-positive Gamma[a] cases and test-negative controls (2-dose analysis)

| | Test Positive (Gamma) N=349 n (%) | Test Negative N=1745 n (%) | p value | ASD |
|---|---|---|---|---|
| Age at specimen collection date, years | | | N/A | N/A |
| 18-44 | 216 (61.9) | 1080 (61.9) | | |
| 45-64 | 97 (27.8) | 485 (27.8) | | |
| 65-74 | 24 (6.9) | 120 (6.9) | | |
| ≥75 | 12 (3.4) | 60 (3.4) | | |
| Sex | | | N/A | N/A |
| Female | 193 (55.3) | 965 (55.3) | | |
| Male | 156 (44.7) | 780 (44.7) | | |
| Race/ethnicity | | | N/A | N/A |
| Non-Hispanic White | 96 (27.5) | 480 (27.5) | | |
| Non-Hispanic Black | 54 (15.5) | 270 (15.5) | | |
| Hispanic | 163 (46.7) | 815 (46.7) | | |
| Non-Hispanic Asian | 7 (2.0) | 35 (2.0) | | |
| Other/Unknown | 29 (8.3) | 145 (8.3) | | |
| BMI[c] | | | 0.1450 | 0.1811 |
| <18.5 | 7 (2.0) | 16 (0.9) | | |
| 18.5 - <25 | 59 (16.9) | 349 (20.0) | | |
| 25 - <30 | 95 (27.2) | 493 (28.3) | | |
| 30 - <35 | 73 (20.9) | 399 (22.9) | | |
| 35 - <40 | 41 (11.7) | 205 (11.7) | | |
| 40 - <45 | 18 (5.2) | 79 (4.5) | | |
| ≥45 | 11 (3.2) | 52 (3.0) | | |
| Unknown | 45 (12.9) | 152 (8.7) | | |
| Smoking[c] | | | 0.0778 | 0.1252 |
| No | 253 (72.5) | 1288 (73.8) | | |
| Yes | 61 (17.5) | 340 (19.5) | | |
| Unknown | 35 (10.0) | 117 (6.7) | | |
| Charlson comorbidity score[b] | | | 0.0038 | 0.2109 |
| 0 | 286 (81.9) | 1302 (74.6) | | |
| 1 | 38 (10.9) | 213 (12.2) | | |
| ≥2 | 25 (7.2) | 230 (13.2) | | |
| Frailty index[b] | | | 0.0039 | 0.2110 |
| Quartile 1 | 84 (24.1) | 421 (24.1) | | |
| Quartile 2 | 116 (33.2) | 426 (24.4) | | |
| Quartile 3 | 77 (22.1) | 447 (25.6) | | |
| Quartile 4 (most frail) | 72 (20.6) | 451 (25.8) | | |
| Chronic diseases[b] | | | | |
| Kidney disease | 8 (2.3) | 64 (3.7) | 0.1980 | 0.0810 |
| Heart disease | 4 (1.1) | 44 (2.5) | 0.1171 | 0.1026 |
| Lung disease | 17 (4.9) | 146 (8.4) | 0.0261 | 0.1410 |
| Liver disease | 5 (1.4) | 65 (3.7) | 0.0296 | 0.1450 |
| Diabetes | 33 (9.5) | 177 (10.1) | 0.6962 | 0.0231 |
| Immunocompromised[e] | 5 (1.4) | 75 (4.3) | 0.0108 | 0.1724 |
| Autoimmune conditions[b,f] | 5 (1.4) | 43 (2.5) | 0.2398 | 0.0747 |
| Pregnant at specimen collection date | 8 (2.3) | 98 (5.6) | 0.0097 | 0.1712 |
| History of COVID-19 diagnosis[d] | 2 (0.6) | 116 (6.6) | <0.0001 | 0.3300 |
| History of SARS-CoV-2 molecular test[d] | 174 (49.9) | 899 (51.5) | 0.5707 | 0.0332 |
| Number of outpatient and virtual visits[b] | | | <0.0001 | 0.3922 |
| 0 | 36 (10.3) | 105 (6.0) | | |
| 1-4 | 132 (37.8) | 439 (25.2) | | |
| 5-10 | 90 (25.8) | 469 (26.9) | | |
| ≥11 | 91 (26.1) | 732 (41.9) | | |

| | | | | |
|---|---|---|---|---|
| Number of ED visits[b] | | | 0.0004 | 0.2555 |
| 0 | 304 (87.1) | 1367 (78.3) | | |
| 1 | 33 (9.5) | 230 (13.2) | | |
| ≥2 | 12 (3.4) | 148 (8.5) | | |
| Number of hospitalizations[b] | | | 0.0335 | 0.1784 |
| 0 | 338 (96.8) | 1632 (93.5) | | |
| 1 | 10 (2.9) | 82 (4.7) | | |
| ≥2 | 1 (0.3) | 31 (1.8) | | |
| Preventive care[b] | 179 (51.3) | 1155 (66.2) | <0.0001 | 0.3062 |
| Medicaid | 41 (11.7) | 189 (10.8) | 0.6170 | 0.0290 |
| Neighborhood median household income | | | 0.4784 | 0.0988 |
| < $40,000 | 17 (4.9) | 101 (5.8) | | |
| $40,000-$59,999 | 69 (19.8) | 390 (22.3) | | |
| $60,000-$79,999 | 94 (26.9) | 434 (24.9) | | |
| $80,000+ | 168 (48.1) | 819 (46.9) | | |
| Unknown | 1 (0.3) | 1 (0.1) | | |
| KPSC physician/employee | 5 (1.4) | 77 (4.4) | 0.0088 | 0.1776 |
| Month of specimen collection | | | 0.1218 | 0.1562 |
| March 2021 | 34 (9.7) | 196 (11.2) | | |
| April 2021 | 106 (30.4) | 459 (26.3) | | |
| May 2021 | 92 (26.4) | 529 (30.3) | | |
| June 2021 | 68 (19.5) | 374 (21.4) | | |
| July 2021 | 49 (14.0) | 187 (10.7) | | |
| Specimen type | | | <0.0001 | 0.4793 |
| NP/OP swab | 340 (97.4) | 1463 (83.8) | | |
| Saliva (asymptomatic) | 9 (2.6) | 282 (16.2) | | |

[a] Gamma variant lineages: P.1, P.1.1
[b] Defined in the one year prior to specimen collection date
[c] Defined in the two years prior to specimen collection date
[d] Defined based on all available medical records from March 1, 2020 to specimen collection date
[e] Defined as HIV/AIDS; leukemia/lymphoma, congenital and other immunodeficiencies, asplenia/hyposplenia; organ transplant; immunosuppressant medications
[f] Defined as rheumatoid arthritis, inflammatory bowel disease, psoriasis and psoriatic arthritis, multiple sclerosis, systemic lupus erythematosus
Medical center area not shown. There were differences in the distribution of cases and controls across the medical center areas.
Abbreviations: ASD, Absolute Standardized Difference; BMI, Body Mass Index; ED, Emergency Department; HIV/AIDS, Human Immunodeficiency Virus/Acquired Immunodeficiency Syndrome; NP/OP, nasopharyngeal/oropharyngeal

### Table S5. Characteristics of SARS-CoV-2 test-positive Iota[a] cases and test-negative controls (2-dose analysis)

| | Test Positive (Iota) N=114 n (%) | Test Negative N=570 n (%) | p value | ASD |
|---|---|---|---|---|
| Age at specimen collection date, years | | | N/A | N/A |
| 18-44 | 68 (59.6) | 340 (59.6) | | |
| 45-64 | 38 (33.3) | 190 (33.3) | | |
| 65-74 | 8 (7.0) | 40 (7.0) | | |
| ≥75 | 0 (0.0) | 0 (0.0) | | |
| Sex | | | N/A | N/A |
| Female | 72 (63.2) | 360 (63.2) | | |
| Male | 42 (36.8) | 210 (36.8) | | |
| Race/ethnicity | | | N/A | N/A |
| Non-Hispanic White | 40 (35.1) | 200 (35.1) | | |
| Non-Hispanic Black | 18 (15.8) | 90 (15.8) | | |
| Hispanic | 37 (32.5) | 185 (32.5) | | |
| Non-Hispanic Asian | 7 (6.1) | 35 (6.1) | | |
| Other/Unknown | 12 (10.5) | 60 (10.5) | | |
| BMI[c] | | | 0.8225 | 0.1984 |
| <18.5 | 1 (0.9) | 6 (1.1) | | |
| 18.5 - <25 | 35 (30.7) | 141 (24.7) | | |
| 25 - <30 | 25 (21.9) | 142 (24.9) | | |
| 30 - <35 | 23 (20.2) | 121 (21.2) | | |
| 35 - <40 | 15 (13.2) | 61 (10.7) | | |
| 40 - <45 | 4 (3.5) | 23 (4.0) | | |
| ≥45 | 4 (3.5) | 21 (3.7) | | |
| Unknown | 7 (6.1) | 55 (9.6) | | |
| Smoking[c] | | | 0.3144 | 0.1636 |
| No | 88 (77.2) | 409 (71.8) | | |
| Yes | 16 (14.0) | 115 (20.2) | | |
| Unknown | 10 (8.8) | 46 (8.1) | | |
| Charlson comorbidity score[b] | | | 0.1049 | 0.2352 |
| 0 | 96 (84.2) | 431 (75.6) | | |
| 1 | 11 (9.6) | 70 (12.3) | | |
| ≥2 | 7 (6.1) | 69 (12.1) | | |
| Frailty index[b] | | | 0.7551 | 0.1127 |
| Quartile 1 | 25 (21.9) | 135 (23.7) | | |
| Quartile 2 | 32 (28.1) | 146 (25.6) | | |
| Quartile 3 | 32 (28.1) | 143 (25.1) | | |
| Quartile 4 (most frail) | 25 (21.9) | 146 (25.6) | | |
| Chronic diseases[b] | | | | |
| Kidney disease | 3 (2.6) | 17 (3.0) | 0.8391 | 0.0212 |
| Heart disease | 2 (1.8) | 14 (2.5) | 0.6509 | 0.0489 |
| Lung disease | 5 (4.4) | 57 (10.0) | 0.0567 | 0.2186 |
| Liver disease | 1 (0.9) | 17 (3.0) | 0.1999 | 0.1535 |
| Diabetes | 3 (2.6) | 55 (9.6) | 0.0141 | 0.2955 |
| Immunocompromised[e] | 3 (2.6) | 25 (4.4) | 0.3881 | 0.0955 |
| Autoimmune conditions[b,f] | 3 (2.6) | 16 (2.8) | 0.9171 | 0.0108 |
| Pregnant at specimen collection date | 2 (1.8) | 31 (5.4) | 0.0938 | 0.1988 |
| History of COVID-19 diagnosis[d] | 1 (0.9) | 44 (7.7) | 0.0071 | 0.3423 |
| History of SARS-CoV-2 molecular test[d] | 59 (51.8) | 298 (52.3) | 0.9182 | 0.0105 |
| Number of outpatient and virtual visits[b] | | | 0.0998 | 0.2551 |
| 0 | 10 (8.8) | 33 (5.8) | | |
| 1-4 | 40 (35.1) | 152 (26.7) | | |
| 5-10 | 28 (24.6) | 147 (25.8) | | |
| ≥11 | 36 (31.6) | 238 (41.8) | | |

| | | | | |
|---|---|---|---|---|
| Number of ED visits[b] | | | 0.3980 | 0.1463 |
| 0 | 98 (86.0) | 460 (80.7) | | |
| 1 | 12 (10.5) | 78 (13.7) | | |
| ≥2 | 4 (3.5) | 32 (5.6) | | |
| Number of hospitalizations[b] | | | 0.1555 | 0.2373 |
| 0 | 112 (98.2) | 535 (93.9) | | |
| 1 | 2 (1.8) | 29 (5.1) | | |
| ≥2 | 0 (0.0) | 6 (1.1) | | |
| Preventive care[b] | 56 (49.1) | 379 (66.5) | 0.0004 | 0.3572 |
| Medicaid | 10 (8.8) | 61 (10.7) | 0.5374 | 0.0651 |
| Neighborhood median household income | | | 0.0475 | 0.2861 |
| < $40,000 | 9 (7.9) | 23 (4.0) | | |
| $40,000-$59,999 | 19 (16.7) | 120 (21.1) | | |
| $60,000-$79,999 | 19 (16.7) | 141 (24.7) | | |
| $80,000+ | 67 (58.8) | 286 (50.2) | | |
| Unknown | 0 (0.0) | 0 (0.0) | | |
| KPSC physician/employee | 9 (7.9) | 24 (4.2) | 0.0938 | 0.1550 |
| Month of specimen collection | | | 0.4537 | 0.1921 |
| March 2021 | 29 (25.4) | 163 (28.6) | | |
| April 2021 | 47 (41.2) | 191 (33.5) | | |
| May 2021 | 24 (21.1) | 149 (26.1) | | |
| June 2021 | 12 (10.5) | 62 (10.9) | | |
| July 2021 | 2 (1.8) | 5 (0.9) | | |
| Specimen type | | | <0.0001 | 0.5208 |
| NP/OP swab | 112 (98.2) | 478 (83.9) | | |
| Saliva (asymptomatic) | 2 (1.8) | 92 (16.1) | | |

[a] Iota variant lineages: B.1.526, B.1.526.1, B.1.526.2
[b] Defined in the one year prior to specimen collection date
[c] Defined in the two years prior to specimen collection date
[d] Defined based on all available medical records from March 1, 2020 to specimen collection date
[e] Defined as HIV/AIDS; leukemia/lymphoma, congenital and other immunodeficiencies, asplenia/hyposplenia; organ transplant; immunosuppressant medications
[f] Defined as rheumatoid arthritis, inflammatory bowel disease, psoriasis and psoriatic arthritis, multiple sclerosis, systemic lupus erythematosus
Medical center area not shown. There were no differences in the distribution of cases and controls across the medical center areas.
Abbreviations: ASD, Absolute Standardized Difference; BMI, Body Mass Index; ED, Emergency Department; HIV/AIDS, Human Immunodeficiency Virus/Acquired Immunodeficiency Syndrome; NP/OP, nasopharyngeal/oropharyngeal

### Table S6. Characteristics of SARS-CoV-2 test-positive Mu[a] cases and test-negative controls (2-dose analysis)

| | Test Positive (Mu) N=69 n (%) | Test Negative N=345 n (%) | p value | ASD |
|---|---|---|---|---|
| Age at specimen collection date, years | | | N/A | N/A |
| 18-44 | 46 (66.7) | 230 (66.7) | | |
| 45-64 | 21 (30.4) | 105 (30.4) | | |
| 65-74 | 2 (2.9) | 10 (2.9) | | |
| ≥75 | 0 (0.0) | 0 (0.0) | | |
| Sex | | | N/A | N/A |
| Female | 37 (53.6) | 185 (53.6) | | |
| Male | 32 (46.4) | 160 (46.4) | | |
| Race/ethnicity | | | N/A | N/A |
| Non-Hispanic White | 23 (33.3) | 115 (33.3) | | |
| Non-Hispanic Black | 7 (10.1) | 35 (10.1) | | |
| Hispanic | 28 (40.6) | 140 (40.6) | | |
| Non-Hispanic Asian | 5 (7.2) | 25 (7.2) | | |
| Other/Unknown | 6 (8.7) | 30 (8.7) | | |
| BMI[c] | | | 0.0949 | 0.4715 |
| <18.5 | 0 (0.0) | 2 (0.6) | | |
| 18.5 - <25 | 21 (30.4) | 77 (22.3) | | |
| 25 - <30 | 12 (17.4) | 99 (28.7) | | |
| 30 - <35 | 20 (29.0) | 55 (15.9) | | |
| 35 - <40 | 6 (8.7) | 40 (11.6) | | |
| 40 - <45 | 3 (4.3) | 18 (5.2) | | |
| ≥45 | 1 (1.4) | 14 (4.1) | | |
| Unknown | 6 (8.7) | 40 (11.6) | | |
| Smoking[c] | | | 0.5787 | 0.1442 |
| No | 54 (78.3) | 253 (73.3) | | |
| Yes | 9 (13.0) | 63 (18.3) | | |
| Unknown | 6 (8.7) | 29 (8.4) | | |
| Charlson comorbidity score[b] | | | 0.2476 | 0.2425 |
| 0 | 57 (82.6) | 262 (75.9) | | |
| 1 | 5 (7.2) | 51 (14.8) | | |
| ≥2 | 7 (10.1) | 32 (9.3) | | |
| Frailty index[b] | | | 0.9769 | 0.0600 |
| Quartile 1 | 18 (26.1) | 86 (24.9) | | |
| Quartile 2 | 17 (24.6) | 86 (24.9) | | |
| Quartile 3 | 18 (26.1) | 85 (24.6) | | |
| Quartile 4 (most frail) | 16 (23.2) | 88 (25.5) | | |
| Chronic diseases[b] | | | | |
| Kidney disease | 0 (0.0) | 9 (2.6) | 0.1750 | 0.2315 |
| Heart disease | 2 (2.9) | 6 (1.7) | 0.5230 | 0.0771 |
| Lung disease | 6 (8.7) | 25 (7.2) | 0.6763 | 0.0535 |
| Liver disease | 1 (1.4) | 14 (4.1) | 0.2898 | 0.1599 |
| Diabetes | 4 (5.8) | 37 (10.7) | 0.2110 | 0.1797 |
| Immunocompromised status | 1 (1.4) | 11 (3.2) | 0.4318 | 0.1157 |
| Autoimmune conditions[b, f] | 3 (4.3) | 5 (1.4) | 0.1103 | 0.1734 |
| Pregnant at specimen collect date | 2 (2.9) | 16 (4.6) | 0.5178 | 0.0914 |
| History of COVID-19 diagnosis[d] | 1 (1.4) | 29 (8.4) | 0.0419 | 0.3256 |
| History of SARS-CoV-2 molecular test[d] | 46 (66.7) | 161 (46.7) | 0.0024 | 0.4121 |
| Number of outpatient and virtual visits[b] | | | 0.6207 | 0.1755 |
| 0 | 6 (8.7) | 32 (9.3) | | |
| 1-4 | 22 (31.9) | 87 (25.2) | | |
| 5-10 | 17 (24.6) | 107 (31.0) | | |
| ≥11 | 24 (34.8) | 119 (34.5) | | |

| | | | | |
|---|---|---|---|---|
| Number of ED visits[b] | | | 0.5225 | 0.1516 |
| 0 | 56 (81.2) | 287 (83.2) | | |
| 1 | 11 (15.9) | 41 (11.9) | | |
| ≥2 | 2 (2.9) | 17 (4.9) | | |
| Number of hospitalizations[b] | | | 0.5620 | 0.1791 |
| 0 | 67 (97.1) | 327 (94.8) | | |
| 1 | 2 (2.9) | 13 (3.8) | | |
| ≥2 | 0 (0.0) | 5 (1.4) | | |
| Preventive care[b] | 41 (59.4) | 211 (61.2) | 0.7870 | 0.0355 |
| Medicaid | 5 (7.2) | 36 (10.4) | 0.4183 | 0.1125 |
| Neighborhood median household income | | | 0.8166 | 0.1755 |
| < $40,000 | 4 (5.8) | 20 (5.8) | | |
| $40,000-$59,999 | 12 (17.4) | 55 (15.9) | | |
| $60,000-$79,999 | 15 (21.7) | 98 (28.4) | | |
| $80,000+ | 38 (55.1) | 171 (49.6) | | |
| Unknown | 0 (0.0) | 1 (0.3) | | |
| KPSC physician/employee | 5 (7.2) | 13 (3.8) | 0.1959 | 0.1529 |
| Month of specimen collection | | | 0.0168 | 0.3668 |
| March 2021 | 0 (0.0) | 0 (0.0) | | |
| April 2021 | 1 (1.4) | 0 (0.0) | | |
| May 2021 | 5 (7.2) | 27 (7.8) | | |
| June 2021 | 12 (17.4) | 107 (31.0) | | |
| July 2021 | 51 (73.9) | 211 (61.2) | | |
| Specimen type | | | 0.0213 | 0.3527 |
| NP/OP swab | 66 (95.7) | 295 (85.5) | | |
| Saliva (asymptomatic) | 3 (4.3) | 50 (14.5) | | |

[a] Mu variant lineages: B.1.621, B.1.621.1

[b] Defined in the one year prior to specimen collection date

[c] Defined in the two years prior to specimen collection date

[d] Defined based on all available medical records from March 1, 2020 to specimen collection date

[e] Defined as HIV/AIDS; leukemia/lymphoma, congenital and other immunodeficiencies, asplenia/hyposplenia; organ transplant; immunosuppressant medications

[f] Defined as rheumatoid arthritis, inflammatory bowel disease, psoriasis and psoriatic arthritis, multiple sclerosis, systemic lupus erythematosus

Medical center area not shown. There were no differences in the distribution of cases and controls across the medical center areas.

Abbreviations: ASD, Absolute Standardized Difference; BMI, Body Mass Index; ED, Emergency Department; HIV/AIDS, Human Immunodeficiency Virus/Acquired Immunodeficiency Syndrome; NP/OP, nasopharyngeal/oropharyngeal

Table S7. Characteristics of SARS-CoV-2 test-positive other[a] variant cases and test-negative controls (2-dose analysis)

| | Test Positive (Beta, Eta, Kappa, and other variants) N=568 n (%) | Test Negative N=2840 n (%) | p value | ASD |
|---|---|---|---|---|
| Age at specimen collection date, years | | | N/A | N/A |
| 18-44 | 324 (57.0) | 1620 (57.0) | | |
| 45-64 | 200 (35.2) | 1000 (35.2) | | |
| 65-74 | 32 (5.6) | 160 (5.6) | | |
| ≥75 | 12 (2.1) | 60 (2.1) | | |
| Sex | | | N/A | N/A |
| Female | 307 (54.0) | 1535 (54.0) | | |
| Male | 261 (46.0) | 1305 (46.0) | | |
| Race/ethnicity | | | N/A | N/A |
| Non-Hispanic White | 162 (28.5) | 810 (28.5) | | |
| Non-Hispanic Black | 53 (9.3) | 265 (9.3) | | |
| Hispanic | 303 (53.3) | 1515 (53.3) | | |
| Non-Hispanic Asian | 20 (3.5) | 100 (3.5) | | |
| Other/Unknown | 30 (5.3) | 150 (5.3) | | |
| BMI[b] | | | 0.0806 | 0.1584 |
| <18.5 | 5 (0.9) | 39 (1.4) | | |
| 18.5 - <25 | 110 (19.4) | 595 (21.0) | | |
| 25 - <30 | 171 (30.1) | 861 (30.3) | | |
| 30 - <35 | 122 (21.5) | 610 (21.5) | | |
| 35 - <40 | 57 (10.0) | 284 (10.0) | | |
| 40 - <45 | 18 (3.2) | 128 (4.5) | | |
| ≥45 | 19 (3.3) | 106 (3.7) | | |
| Unknown | 66 (11.6) | 217 (7.6) | | |
| Smoking[b] | | | 0.0095 | 0.1352 |
| No | 426 (75.0) | 2121 (74.7) | | |
| Yes | 91 (16.0) | 549 (19.3) | | |
| Unknown | 51 (9.0) | 170 (6.0) | | |
| Charlson comorbidity score[a] | | | 0.0052 | 0.1593 |
| 0 | 449 (79.0) | 2122 (74.7) | | |
| 1 | 75 (13.2) | 362 (12.7) | | |
| ≥2 | 44 (7.7) | 356 (12.5) | | |
| Frailty index[a] | | | 0.0007 | 0.1892 |
| Quartile 1 | 141 (24.8) | 711 (25.0) | | |
| Quartile 2 | 176 (31.0) | 676 (23.8) | | |
| Quartile 3 | 138 (24.3) | 714 (25.1) | | |
| Quartile 4 (most frail) | 113 (19.9) | 739 (26.0) | | |
| Chronic diseases[a] | | | | |
| Kidney disease | 11 (1.9) | 111 (3.9) | 0.0209 | 0.1173 |
| Heart disease | 7 (1.2) | 88 (3.1) | 0.0136 | 0.1285 |
| Lung disease | 37 (6.5) | 234 (8.2) | 0.1653 | 0.0660 |
| Liver disease | 16 (2.8) | 116 (4.1) | 0.1529 | 0.0695 |
| Diabetes | 49 (8.6) | 318 (11.2) | 0.0712 | 0.0861 |
| Immunocompromised[d] | 16 (2.8) | 125 (4.4) | 0.0835 | 0.0850 |
| Autoimmune conditions[a,e] | 14 (2.5) | 103 (3.6) | 0.1650 | 0.0677 |
| Pregnant at specimen collection date | 11 (1.9) | 152 (5.4) | 0.0005 | 0.1830 |
| History of COVID-19 diagnosis[c] | 4 (0.7) | 225 (7.9) | <0.0001 | 0.3610 |
| History of SARS-CoV-2 molecular test[c] | 311 (54.8) | 1541 (54.3) | 0.8295 | 0.0099 |
| Number of outpatient and virtual visits[a] | | | <0.0001 | 0.3373 |
| 0 | 56 (9.9) | 193 (6.8) | | |
| 1-4 | 197 (34.7) | 730 (25.7) | | |

| | | | | |
|---|---|---|---|---|
| 5-10 | 169 (29.8) | 756 (26.6) | | |
| ≥11 | 146 (25.7) | 1161 (40.9) | | |
| Number of ED visits[a] | | | <0.0001 | 0.2350 |
| 0 | 489 (86.1) | 2215 (78.0) | | |
| 1 | 60 (10.6) | 403 (14.2) | | |
| ≥2 | 19 (3.3) | 222 (7.8) | | |
| Number of hospitalizations[a] | | | 0.0003 | 0.2174 |
| 0 | 548 (96.5) | 2615 (92.1) | | |
| 1 | 19 (3.3) | 169 (6.0) | | |
| ≥2 | 1 (0.2) | 56 (2.0) | | |
| Preventive care[a] | 309 (54.4) | 1867 (65.7) | <0.0001 | 0.2331 |
| Medicaid | 44 (7.7) | 303 (10.7) | 0.0355 | 0.1012 |
| Neighborhood median household income | | | 0.7262 | 0.0699 |
| < $40,000 | 27 (4.8) | 147 (5.2) | | |
| $40,000-$59,999 | 123 (21.7) | 645 (22.7) | | |
| $60,000-$79,999 | 137 (24.1) | 725 (25.5) | | |
| $80,000+ | 281 (49.5) | 1321 (46.5) | | |
| Unknown | 0 (0.0) | 2 (0.1) | | |
| KPSC physician/employee | 18 (3.2) | 104 (3.7) | 0.5638 | 0.0271 |
| Month of specimen collection | | | 0.0003 | 0.1943 |
| March 2021 | 284 (50.0) | 1496 (52.7) | | |
| April 2021 | 193 (34.0) | 835 (29.4) | | |
| May 2021 | 53 (9.3) | 323 (11.4) | | |
| June 2021 | 20 (3.5) | 152 (5.4) | | |
| July 2021 | 18 (3.2) | 34 (1.2) | | |
| Specimen type | | | <0.0001 | 0.4570 |
| NP/OP swab | 550 (96.8) | 2374 (83.6) | | |
| Saliva (asymptomatic) | 18 (3.2) | 466 (16.4) | | |

[a] Other variant lineages: A, A.2, A.2.5, A.2.5.1, A.5, B, B.1, B.1.1, B.1.1.174, B.1.1.222, B.1.1.28, B.1.1.304, B.1.1.316, B.1.1.318, B.1.1.328, B.1.1.335, B.1.1.368, B.1.1.378, B.1.1.413, B.1.1.416, B.1.1.432, B.1.1.519, B.1.126, B.1.153, B.1.160, B.1.2, B.1.232, B.1.234, B.1.241, B.1.243, B.1.280, B.1.289, B.1.311, B.1.324, B.1.349, B.1.351 (Beta), B.1.351.3 (Beta), B.1.36.21, B.1.377, B.1.384, B.1.393, B.1.396, B.1.400, B.1.404, B.1.438.1, B.1.441, B.1.523, B.1.525 (Eta), B.1.530, B.1.550, B.1.551, B.1.555, B.1.561, B.1.575, B.1.577, B.1.595, B.1.596, B.1.599, B.1.604, B.1.609, B.1.612, B.1.617.1 (Kappa), B.1.619, B.1.623, B.1.628, C.36, C.37 (Lambda), D.2, P.2 (Zeta), R.1

[b] Defined in the one year prior to specimen collection date

[c] Defined in the two years prior to specimen collection date

[d] Defined based on all available medical records from March 1, 2020 to specimen collection date

[e] Defined as HIV/AIDS; leukemia/lymphoma, congenital and other immunodeficiencies, asplenia/hyposplenia; organ transplant; immunosuppressant medications

[f] Defined as rheumatoid arthritis, inflammatory bowel disease, psoriasis and psoriatic arthritis, multiple sclerosis, systemic lupus erythematosus

Medical center area not shown. There were differences in the distribution of cases and controls across the medical center areas.

Abbreviations: ASD, Absolute Standardized Difference; BMI, Body Mass Index; ED, Emergency Department; HIV/AIDS, Human Immunodeficiency Virus/Acquired Immunodeficiency Syndrome; NP/OP, nasopharyngeal/oropharyngeal

Table S8. Characteristics of SARS-CoV-2 test-positive unidentified variants cases and test-negative controls (2-dose analysis)

| | Test Positive (Unidentified variants) N=2909 n (%) | Test Negative N=14545 n (%) | p value | ASD |
|---|---|---|---|---|
| Age at specimen collection date, years | | | N/A | N/A |
| 18-44 | 1784 (61.3) | 8920 (61.3) | | |
| 45-64 | 869 (29.9) | 4345 (29.9) | | |
| 65-74 | 179 (6.2) | 895 (6.2) | | |
| ≥75 | 77 (2.6) | 385 (2.6) | | |
| Sex | | | N/A | N/A |
| Female | 1618 (55.6) | 8090 (55.6) | | |
| Male | 1291 (44.4) | 6455 (44.4) | | |
| Race/ethnicity | | | N/A | N/A |
| Non-Hispanic White | 786 (27.0) | 3930 (27.0) | | |
| Non-Hispanic Black | 427 (14.7) | 2135 (14.7) | | |
| Hispanic | 1333 (45.8) | 6665 (45.8) | | |
| Non-Hispanic Asian | 155 (5.3) | 775 (5.3) | | |
| Other/Unknown | 208 (7.2) | 1040 (7.2) | | |
| BMI[b] | | | <0.0001 | 0.1378 |
| <18.5 | 29 (1.0) | 172 (1.2) | | |
| 18.5 - <25 | 600 (20.6) | 3120 (21.5) | | |
| 25 - <30 | 803 (27.6) | 4099 (28.2) | | |
| 30 - <35 | 575 (19.8) | 3118 (21.4) | | |
| 35 - <40 | 306 (10.5) | 1563 (10.7) | | |
| 40 - <45 | 132 (4.5) | 721 (5.0) | | |
| ≥45 | 85 (2.9) | 467 (3.2) | | |
| Unknown | 379 (13.0) | 1285 (8.8) | | |
| Smoking[b] | | | <0.0001 | 0.1533 |
| No | 2143 (73.7) | 10811 (74.3) | | |
| Yes | 454 (15.6) | 2740 (18.8) | | |
| Unknown | 312 (10.7) | 994 (6.8) | | |
| Charlson comorbidity score[a] | | | <0.0001 | 0.1134 |
| 0 | 2321 (79.8) | 10962 (75.4) | | |
| 1 | 324 (11.1) | 1822 (12.5) | | |
| ≥2 | 264 (9.1) | 1761 (12.1) | | |
| Frailty index[a] | | | <0.0001 | 0.1763 |
| Quartile 1 | 690 (23.7) | 3626 (24.9) | | |
| Quartile 2 | 912 (31.4) | 3499 (24.1) | | |
| Quartile 3 | 700 (24.1) | 3664 (25.2) | | |
| Quartile 4 (most frail) | 607 (20.9) | 3756 (25.8) | | |
| Chronic diseases[a] | | | | |
| Kidney disease | 86 (3.0) | 487 (3.3) | 0.2789 | 0.0224 |
| Heart disease | 57 (2.0) | 362 (2.5) | 0.0886 | 0.0359 |
| Lung disease | 221 (7.6) | 1215 (8.4) | 0.1754 | 0.0279 |
| Liver disease | 55 (1.9) | 498 (3.4) | <0.0001 | 0.0954 |
| Diabetes | 247 (8.5) | 1532 (10.5) | 0.0009 | 0.0696 |
| Immunocompromised[d] | 87 (3.0) | 644 (4.4) | 0.0004 | 0.0761 |
| Autoimmune conditions[a.e] | 51 (1.8) | 406 (2.8) | 0.0014 | 0.0697 |
| Pregnant at specimen collection date | 74 (2.5) | 742 (5.1) | <0.0001 | 0.1337 |
| History of COVID-19 diagnosis[c] | 94 (3.2) | 1100 (7.6) | <0.0001 | 0.1926 |
| History of SARS-CoV-2 molecular test[c] | 1474 (50.7) | 7554 (51.9) | 0.2126 | 0.0253 |
| Number of outpatient and virtual visits[a] | | | <0.0001 | 0.2619 |
| 0 | 323 (11.1) | 963 (6.6) | | |
| 1-4 | 960 (33.0) | 3908 (26.9) | | |
| 5-10 | 781 (26.8) | 3920 (27.0) | | |

| | | | | |
|---|---|---|---|---|
| ≥11 | 845 (29.0) | 5754 (39.6) | | |
| Number of ED visits[a] | | | <0.0001 | 0.1321 |
| 0 | 2439 (83.8) | 11475 (78.9) | | |
| 1 | 325 (11.2) | 2004 (13.8) | | |
| ≥2 | 145 (5.0) | 1066 (7.3) | | |
| Number of hospitalizations[a] | | | 0.0003 | 0.0919 |
| 0 | 2764 (95.0) | 13660 (93.9) | | |
| 1 | 126 (4.3) | 649 (4.5) | | |
| ≥2 | 19 (0.7) | 236 (1.6) | | |
| Preventive care[a] | 1517 (52.1) | 9568 (65.8) | <0.0001 | 0.2799 |
| Medicaid | 306 (10.5) | 1485 (10.2) | 0.6157 | 0.0102 |
| Neighborhood median household income | | | 0.0342 | 0.0654 |
| < $40,000 | 163 (5.6) | 806 (5.5) | | |
| $40,000-$59,999 | 657 (22.6) | 3175 (21.8) | | |
| $60,000-$79,999 | 797 (27.4) | 3657 (25.1) | | |
| $80,000+ | 1289 (44.3) | 6893 (47.4) | | |
| Unknown | 3 (0.1) | 14 (0.1) | | |
| KPSC physician/employee | 100 (3.4) | 649 (4.5) | 0.0128 | 0.0526 |
| Month of specimen collection | | | <0.0001 | 0.1860 |
| March 2021 | 608 (20.9) | 3142 (21.6) | | |
| April 2021 | 499 (17.2) | 2156 (14.8) | | |
| May 2021 | 359 (12.3) | 2124 (14.6) | | |
| June 2021 | 469 (16.1) | 3122 (21.5) | | |
| July 2021 | 974 (33.5) | 4001 (27.5) | | |
| Specimen type | | | <0.0001 | 0.2096 |
| NP/OP swab | 2644 (90.9) | 12215 (84.0) | | |
| Saliva (asymptomatic) | 265 (9.1) | 2330 (16.0) | | |

[a] Defined in the one year prior to specimen collection date
[b] Defined in the two years prior to specimen collection date
[c] Defined based on all available medical records from March 1, 2020 to specimen collection date
[d] Defined as HIV/AIDS; leukemia/lymphoma, congenital and other immunodeficiencies, asplenia/hyposplenia; organ transplant; immunosuppressant medications
[e] Defined as rheumatoid arthritis, inflammatory bowel disease, psoriasis and psoriatic arthritis, multiple sclerosis, systemic lupus erythematosus
Medical center area not shown. There were differences in the distribution of cases and controls across the medical center areas.
Abbreviations: ASD, Absolute Standardized Difference; BMI, Body Mass Index; ED, Emergency Department; HIV/AIDS, Human Immunodeficiency Virus/Acquired Immunodeficiency Syndrome; NP/OP, nasopharyngeal/oropharyngeal

### Table S9. Vaccine effectiveness of 2 doses of mRNA-1273 against infection with SARS-CoV-2 variants

| | Test Positive | | Test Negative | | Odds Ratio (95% CI) | | VE (95% CI) | |
|---|---|---|---|---|---|---|---|---|
| | Vaccinated n (%) | Unvaccinated n (%) | Vaccinated n (%) | Unvaccinated n (%) | Unadjusted | Adjusted[a] | Unadjusted (%) | Adjusted[a] (%) |
| Alpha | 13 (0.9) | 1409 (99.1) | 1734 (24.4) | 5376 (75.6%) | 0.019 (0.010, 0.034) | 0.016 (0.009, 0.031) | 98.1 (96.6, 99.0) | 98.4 (96.9, 99.1) |
| Delta | 232 (11.4) | 1795 (88.6) | 4588 (45.3) | 5547 (54.7%) | 0.134 (0.115, 0.156) | 0.133 (0.113, 0.157) | 86.6 (84.4, 88.5) | 86.7 (84.3, 88.7) |
| Epsilon | 3 (0.5) | 580 (99.5) | 317 (10.9) | 2598 (89.1%) | 0.024 (0.006, 0.097) | 0.024 (0.006, 0.098) | 97.6 (90.3, 99.4) | 97.6 (90.2, 99.4) |
| Gamma | 9 (2.6) | 340 (97.4) | 552 (31.6) | 1193 (68.4%) | 0.044 (0.022, 0.087) | 0.045 (0.022, 0.091) | 95.6 (91.3, 97.8) | 95.5 (90.9, 97.8) |
| Iota | 3 (2.6) | 111 (97.4) | 122 (21.4) | 448 (78.6%) | 0.076 (0.023, 0.252) | 0.043 (0.010, 0.183) | 92.4 (74.8, 97.7) | 95.7 (81.7, 99.0) |
| Mu | 7 (10.1) | 62 (89.9) | 153 (44.3) | 192 (55.7%) | 0.111 (0.046, 0.267) | 0.096 (0.035, 0.261) | 88.9 (73.3, 95.4) | 90.4 (73.9, 96.5) |
| Other | 6 (1.1) | 562 (98.9) | 457 (16.1) | 2383 (83.9%) | 0.041 (0.018, 0.094) | 0.036 (0.015, 0.088) | 95.9 (90.6, 98.2) | 96.4 (91.2, 98.5) |
| Unidentified | 326 (11.2) | 2583 (88.8) | 4856 (33.4) | 9689 (66.6%) | 0.193 (0.170, 0.220) | 0.201 (0.175, 0.231) | 80.7 (78.0, 83.0) | 79.9 (76.9, 82.5) |

[a] Model adjustment:

Model for Alpha variant adjusted for covariates: BMI, smoking, Charlson comorbidity score, frailty index, lung disease, immunocompromised status, pregnancy, history of COVID-19 diagnosis, number of outpatient and virtual visits, number of ED visits, number of hospitalizations, preventive care, medical center area, month of specimen collection, specimen type.

Model for Delta variant adjusted for covariates: smoking, Charlson comorbidity score, frailty index, liver disease, pregnancy, history of COVID-19 diagnosis, number of outpatient and virtual visits, preventive care, medical center area, month of specimen collection, specimen type.

Model for Epsilon variant adjusted for covariates: BMI, smoking, Charlson comorbidity score, frailty index, kidney disease, heart disease, lung disease, diabetes, immunocompromised status, pregnancy, history of COVID-19 diagnosis, history of SARS-CoV-2 molecular test, number of outpatient and virtual visits, number of ED visits, number of hospitalizations, preventive care, medical center area, specimen type.

Model for Gamma variant adjusted for covariates: smoking, Charlson comorbidity score, frailty index, lung disease, liver disease, immunocompromised status, pregnancy, history of COVID-19 diagnosis, number of outpatient and virtual visits, number of ED visits, number of hospitalizations, preventive care, KPSC physician/employee status, medical center area, specimen type.

Model for Iota variant adjusted for covariates: lung disease, diabetes, pregnancy, history of COVID-19 diagnosis, number of outpatient and virtual visits, preventive care, neighborhood median household income, KPSC physician/employee status, specimen type.

Model for Mu variant adjusted for covariates: BMI, history of COVID-19 diagnosis, history of SARS-CoV-2 molecular test, month of specimen collection, specimen type.

Model for Other variants adjusted for covariates: BMI, smoking, Charlson comorbidity score, frailty index, kidney disease, heart disease, pregnancy, history of COVID-19 diagnosis, number of outpatient and virtual visits, number of ED visits, number of hospitalizations, preventive care, Medicaid, medical center area, month of specimen collection, specimen type.

Model for Unidentified variants adjusted for covariates: BMI, smoking, Charlson comorbidity score, frailty index, pregnancy, history of COVID-19 diagnosis, number of outpatient and virtual visits, number of ED visits, preventive care, medical center area, month of specimen collection, specimen type.

Abbreviations: BMI, Body Mass Index; CI, Confidence Interval; ED, Emergency Department

### Table S10. Vaccine effectiveness of 1 dose of mRNA-1273 against infection with SARS-CoV-2 variants

| | Test Positive | | Test Negative | | Odds Ratio (95% CI) | | VE (95% CI) | |
|---|---|---|---|---|---|---|---|---|
| | Vaccinated n (%) | Unvaccinated n (%) | Vaccinated n (%) | Unvaccinated n (%) | Unadjusted | Adjusted[a] | Unadjusted | Adjusted[a] |
| Alpha | 14 (1.0) | 1409 (99.0) | 635 (8.9) | 6480 (91.1) | 0.098 (0.057, 0.167) | 0.099 (0.058, 0.171) | 90.2 (83.3, 94.3) | 90.1 (82.9, 94.2) |
| Delta | 15 (0.8) | 1795 (99.2) | 336 (3.7) | 8714 (96.3) | 0.215 (0.128, 0.362) | 0.230 (0.135, 0.393) | 78.5 (63.8, 87.2) | 77.0 (60.7, 86.5) |
| Epsilon | 7 (1.2) | 580 (98.8) | 149 (5.1) | 2786 (94.9) | 0.212 (0.098, 0.459) | 0.237 (0.109, 0.519) | 78.8 (54.1, 90.2) | 76.3 (48.1, 89.1) |
| Gamma | 8 (2.3) | 340 (97.7) | 132 (7.6) | 1608 (92.4) | 0.275 (0.132, 0.572) | 0.258 (0.119, 0.562) | 72.5 (42.8, 86.8) | 74.2 (43.8, 88.1) |
| Iota | 1 (0.9) | 111 (99.1) | 29 (5.2) | 531 (94.8) | 0.164 (0.022, 1.216) | 0.112 (0.013, 0.993) | 83.6 (0.0, 97.8) | 88.8 (0.7, 98.7) |
| Mu | 2 (3.1) | 62 (96.9) | 16 (5.0) | 304 (95.0) | 0.614 (0.138, 2.732) | 0.542 (0.111, 2.644) | 38.6 (0.0, 86.2) | 45.8 (0.0, 88.9) |
| Other | 7 (1.2) | 562 (98.8) | 205 (7.2) | 2640 (92.8) | 0.155 (0.072, 0.333) | 0.157 (0.073, 0.341) | 84.5 (66.7, 92.8) | 84.3 (65.9, 92.7) |
| Unidentified | 58 (2.2) | 2583 (97.8) | 828 (6.3) | 12377 (93.7) | 0.326 (0.248, 0.428) | 0.324 (0.244, 0.429) | 67.4 (57.2, 75.2) | 67.6 (57.1, 75.6) |

[a] Model adjustment:

Model for Alpha variant adjusted for covariates: BMI, smoking, Charlson comorbidity score, frailty index, liver disease, immunocompromised status, pregnancy, history of COVID-19 diagnosis, number of outpatient and virtual visits, number of ED visits, number of hospitalizations, preventive care, medical center area, specimen type.

Model for Delta variant adjusted for covariates: Charlson comorbidity score, frailty index, liver disease, pregnancy, history of COVID-19 diagnosis, number of outpatient and virtual visits, number of ED visits, preventive care, medical center area, month of specimen collection, specimen type.

Model for Epsilon variant adjusted for covariates: BMI, smoking, Charlson comorbidity score, frailty index, heart disease, lung disease, liver disease, pregnancy, history of COVID-19 diagnosis, number of outpatient and virtual visits, number of ED visits, number of hospitalizations, preventive care, medical center area, specimen type.

Model for Gamma variant adjusted for covariates: Charlson comorbidity score, frailty index, heart disease, lung disease, liver disease, immunocompromised status, pregnancy, history of COVID-19 diagnosis, number of outpatient and virtual visits, number of ED visits, number of hospitalizations, preventive care, KPSC physician/employee status, medical center area, specimen type.

Model for Iota variant adjusted for covariates: Charlson comorbidity score, lung disease, diabetes, pregnancy, history of COVID-19 diagnosis, number of outpatient and virtual visits, preventive care, KPSC physician/employee status, medical center area, specimen type.

Model for Mu variant adjusted for covariates: BMI, history of COVID-19 diagnosis, history of SARS-CoV-2 molecular test, month of specimen collection, specimen type.

Model for Other variants adjusted for covariates: smoking, Charlson comorbidity score, frailty index, kidney disease, pregnancy, history of COVID-19 diagnosis, number of outpatient and virtual visits, number of ED visits, number of hospitalizations, preventive care, Medicaid, medical center area, month of specimen collection, specimen type.

Model for Unidentified variants adjusted for covariates: BMI, smoking, Charlson comorbidity score, frailty index, liver disease, pregnancy, history of COVID-19 diagnosis, number of outpatient and virtual visits, number of ED visits, preventive care, medical center area, month of specimen collection, specimen type.

Abbreviations: BMI, Body Mass Index; CI, Confidence Interval; ED, Emergency Department

### Table S11. Vaccine effectiveness of 2 doses of mRNA-1273 against infection with SARS-CoV-2 variants by time since vaccination

| | Test Positive | | Test Negative | | Odds Ratio (95% CI) | | VE (95% CI) | |
|---|---|---|---|---|---|---|---|---|
| | Vaccinated n (%) | Unvaccinated n (%) | Vaccinated n (%) | Unvaccinated n (%) | Unadjusted | Adjusted[a] | Unadjusted (%) | Adjusted[a] (%) |
| Delta | | | | | | | | |
| 14-60 days | 21 (1.0) | 1795 (88.6) | 977 (9.6) | 5547 (54.7) | 0.060 (0.039, 0.093) | 0.059 (0.037, 0.095) | 94.0 (90.7, 96.1) | 94.1 (90.5, 96.3) |
| 61-90 days | 58 (2.9) | 1795 (88.6) | 1465 (14.5) | 5547 (54.7) | 0.110 (0.084, 0.145) | 0.113 (0.085, 0.150) | 89.0 (85.5, 91.6) | 88.7 (85.0, 91.5) |
| 91-120 days | 56 (2.8) | 1795 (88.6) | 1023 (10.1) | 5547 (54.7) | 0.141 (0.106, 0.187) | 0.141 (0.105, 0.189) | 85.9 (81.3, 89.4) | 85.9 (81.1, 89.5) |
| 121-150 days | 66 (3.3) | 1795 (88.6) | 710 (7.0) | 5547 (54.7) | 0.232 (0.177, 0.304) | 0.230 (0.171, 0.309) | 76.8 (69.6, 82.3) | 77.0 (69.1, 82.9) |
| 151-180 days | 31 (1.5) | 1795 (88.6) | 395 (3.9) | 5547 (54.7) | 0.217 (0.149, 0.316) | 0.200 (0.134, 0.298) | 78.3 (68.4, 85.1) | 80.0 (70.2, 86.6) |
| >180 days | 0 (0.0) | 1795 (88.6) | 18 (0.2) | 5547 (54.7) | N/E | N/E | N/E | N/E |
| Non-Delta | | | | | | | | |
| 14-60 days | 11 (0.4) | 3064 (98.7) | 2107 (13.6) | 12190 (78.5) | 0.014 (0.008, 0.027) | 0.014 (0.007, 0.027) | 98.6 (97.3, 99.2) | 98.6 (97.3, 99.3) |
| 61-90 days | 10 (0.3) | 3064 (98.7) | 737 (4.7) | 12190 (78.5) | 0.031 (0.016, 0.061) | 0.031 (0.015, 0.061) | 96.9 (93.9, 98.4) | 96.9 (93.9, 98.5) |
| 91-120 days | 12 (0.4) | 3064 (98.7) | 321 (2.1) | 12190 (78.5) | 0.104 (0.058, 0.187) | 0.086 (0.046, 0.161) | 89.6 (81.3, 94.2) | 91.4 (83.9, 95.4) |
| 121-150 days | 8 (0.3) | 3064 (98.7) | 127 (0.8) | 12190 (78.5) | 0.153 (0.072, 0.325) | 0.113 (0.048, 0.268) | 84.7 (67.5, 92.8) | 88.7 (73.2, 95.2) |
| 151-180 days | 0 (0.0) | 3064 (98.7) | 41 (0.3) | 12190 (78.5) | N/E | N/E | N/E | N/E |
| >180 days | 0 (0.0) | 3064 (98.7) | 2 (0.0) | 12190 (78.5) | N/E | N/E | N/E | N/E |
| Unidentified | | | | | | | | |
| 14-60 days | 97 (3.3) | 2583 (88.8) | 2027 (13.9) | 9689 (66.6) | 0.150 (0.121, 0.186) | 0.164 (0.131, 0.205) | 85.0 (81.4, 87.9) | 83.6 (79.5, 86.9) |
| 61-90 days | 76 (2.6) | 2583 (88.8) | 1267 (8.7) | 9689 (66.6) | 0.167 (0.131, 0.214) | 0.178 (0.138, 0.230) | 83.3 (78.6, 86.9) | 82.2 (77.0, 86.2) |
| 91-120 days | 68 (2.3) | 2583 (88.8) | 867 (6.0) | 9689 (66.6) | 0.218 (0.168, 0.284) | 0.223 (0.170, 0.293) | 78.2 (71.6, 83.2) | 77.7 (70.7, 83.0) |
| 121-150 days | 59 (2.0) | 2583 (88.8) | 454 (3.1) | 9689 (66.6) | 0.355 (0.264, 0.477) | 0.336 (0.244, 0.464) | 64.5 (52.3, 73.6) | 66.4 (53.6, 75.6) |
| 151-180 days | 26 (0.9) | 2583 (88.8) | 232 (1.6) | 9689 (66.6) | 0.317 (0.208, 0.483) | 0.315 (0.204, 0.487) | 68.3 (51.7, 79.2) | 68.5 (51.3, 79.6) |
| >180 days | 0 (0.0) | 2583 (88.8) | 9 (0.1) | 9689 (66.6) | N/E | N/E | N/E | N/E |

a Model adjustment:

Model for Delta adjusted for covariates: smoking, Charlson comorbidity score, frailty index, liver disease, pregnancy, history of COVID-19 diagnosis, number of outpatient and virtual visits, preventive care, medical center area, month of specimen collection, specimen type.

Model for Non-Delta variant adjusted for covariates: BMI, smoking, Charlson comorbidity score, frailty index, kidney disease, heart disease, lung disease, immunocompromised status, pregnancy, history of COVID-19 diagnosis, number of outpatient and virtual visits, number of ED visits, number of hospitalizations, preventive care, medical center area, month of specimen collection, specimen type.

Model for Unidentified variants adjusted for covariates: BMI, smoking, Charlson comorbidity score, frailty index, pregnancy, history of COVID-19 diagnosis, number of outpatient and virtual visits, number of ED visits, preventive care, medical center area, month of specimen collection, specimen type.

Abbreviations: BMI, Body Mass Index; CI, Confidence Interval; ED, Emergency Department; N/E, Not Estimable

### Figure S1. Distribution of SARS-CoV-2 variants by mRNA-1273 vaccination status

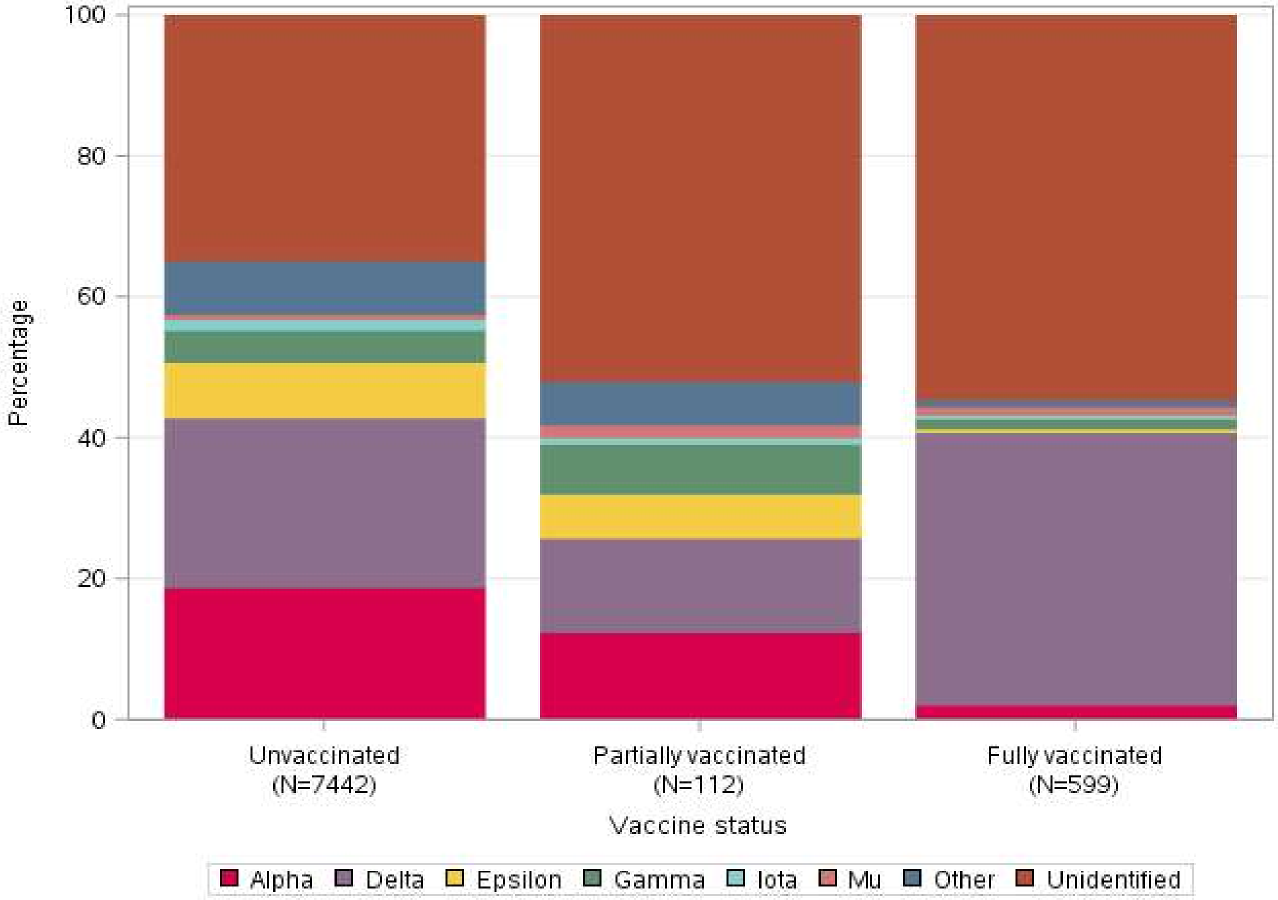

Variant lineages:

Alpha: B.1.1.7

Delta: AY.1, AY.2, AY.3, B.1.617.2

Epsilon: B.1.427, B.1.429

Gamma: P.1, P.1.1

Iota: B.1.526, B.1.526.1, B.1.526.2

Mu: B.1.621, B.1.621.1

Other (Beta, Eta, Kappa, and other variants): A, A.2, A.2.5, A.2.5.1, A.5, B, B.1, B.1.1, B.1.1.174, B.1.1.222, B.1.1.28, B.1.1.304, B.1.1.316, B.1.1.318, B.1.1.328, B.1.1.335, B.1.1.368, B.1.1.378, B.1.1.413, B.1.1.416, B.1.1.432, B.1.1.519, B.1.126, B.1.153, B.1.160, B.1.2, B.1.232, B.1.234, B.1.241, B.1.243, B.1.280, B.1.289, B.1.311, B.1.324, B.1.349, B.1.351 (Beta), B.1.351.3 (Beta), B.1.36.21, B.1.377, B.1.384, B.1.393, B.1.396, B.1.400, B.1.404, B.1.438.1, B.1.441, B.1.523, B.1.525 (Eta), B.1.530, B.1.550, B.1.551, B.1.555, B.1.561, B.1.575, B.1.577, B.1.595, B.1.596, B.1.599, B.1.604, B.1.609, B.1.612, B.1.617.1 (Kappa), B.1.619, B.1.623, B.1.628, C.36, C.37 (Lambda), D.2, P.2 (Zeta), R.1

Figure S2. Distribution of SARS-CoV-2 variants by month of specimen collection

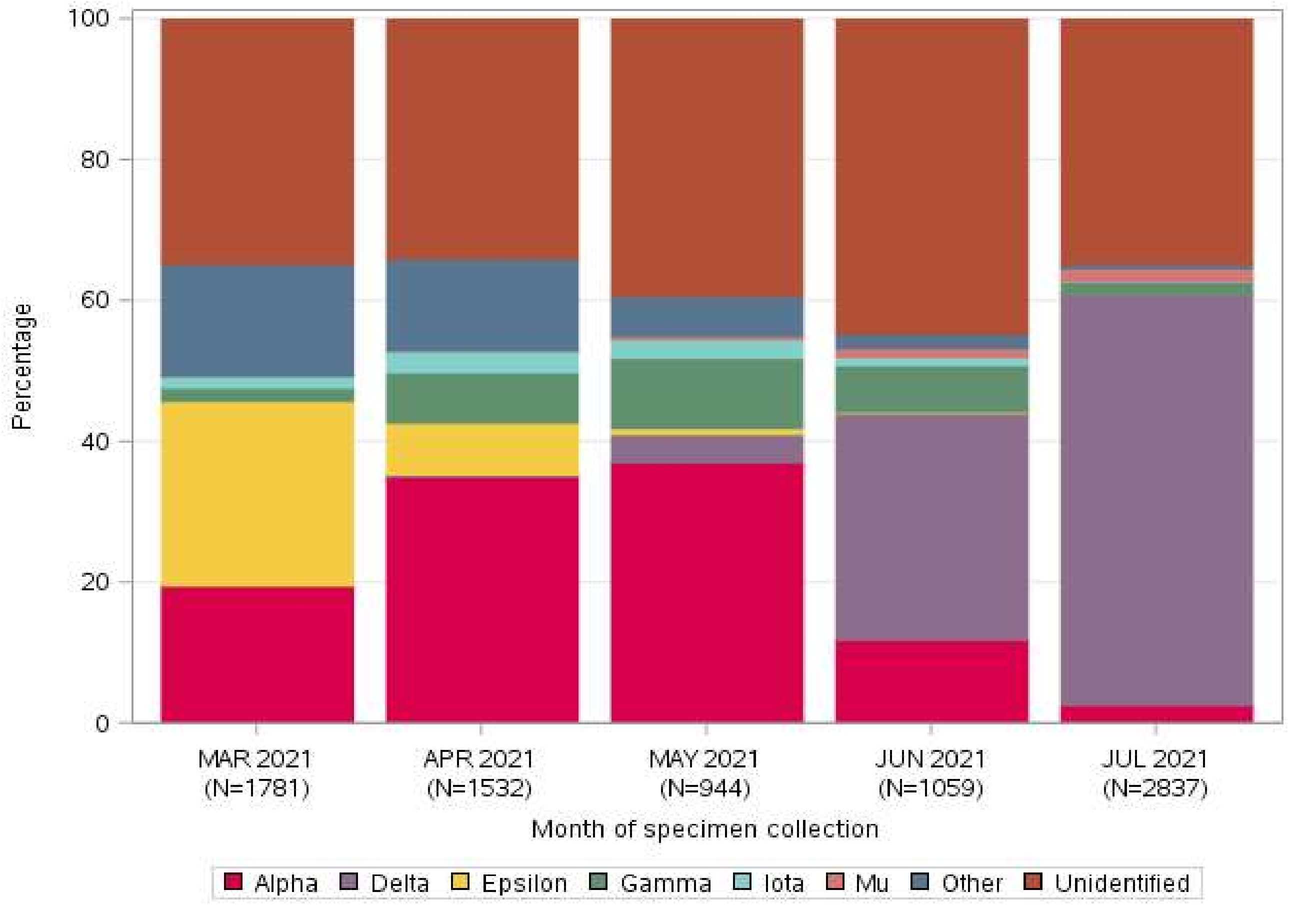

Variant lineages:

Alpha: B.1.1.7

Delta: AY.1, AY.2, AY.3, B.1.617.2

Epsilon: B.1.427, B.1.429

Gamma: P.1, P.1.1

Iota: B.1.526, B.1.526.1, B.1.526.2

Mu: B.1.621, B.1.621.1

Other (Beta, Eta, Kappa, and other variants): A, A.2, A.2.5, A.2.5.1, A.5, B, B.1, B.1.1, B.1.1.174, B.1.1.222, B.1.1.28, B.1.1.304, B.1.1.316, B.1.1.318, B.1.1.328, B.1.1.335, B.1.1.368, B.1.1.378, B.1.1.413, B.1.1.416, B.1.1.432, B.1.1.519, B.1.126, B.1.153, B.1.160, B.1.2, B.1.232, B.1.234, B.1.241, B.1.243, B.1.280, B.1.289, B.1.311, B.1.324, B.1.349, B.1.351 (Beta), B.1.351.3 (Beta), B.1.36.21, B.1.377, B.1.384, B.1.393, B.1.396, B.1.400, B.1.404, B.1.438.1, B.1.441, B.1.523, B.1.525 (Eta), B.1.530, B.1.550, B.1.551, B.1.555, B.1.561, B.1.575, B.1.577, B.1.595, B.1.596, B.1.599, B.1.604, B.1.609, B.1.612, B.1.617.1 (Kappa), B.1.619, B.1.623, B.1.628, C.36, C.37 (Lambda), D.2, P.2 (Zeta), R.1

Figure S3. Vaccine effectiveness of 2 doses of mRNA-1273 against Delta infection by time since vaccination and age group

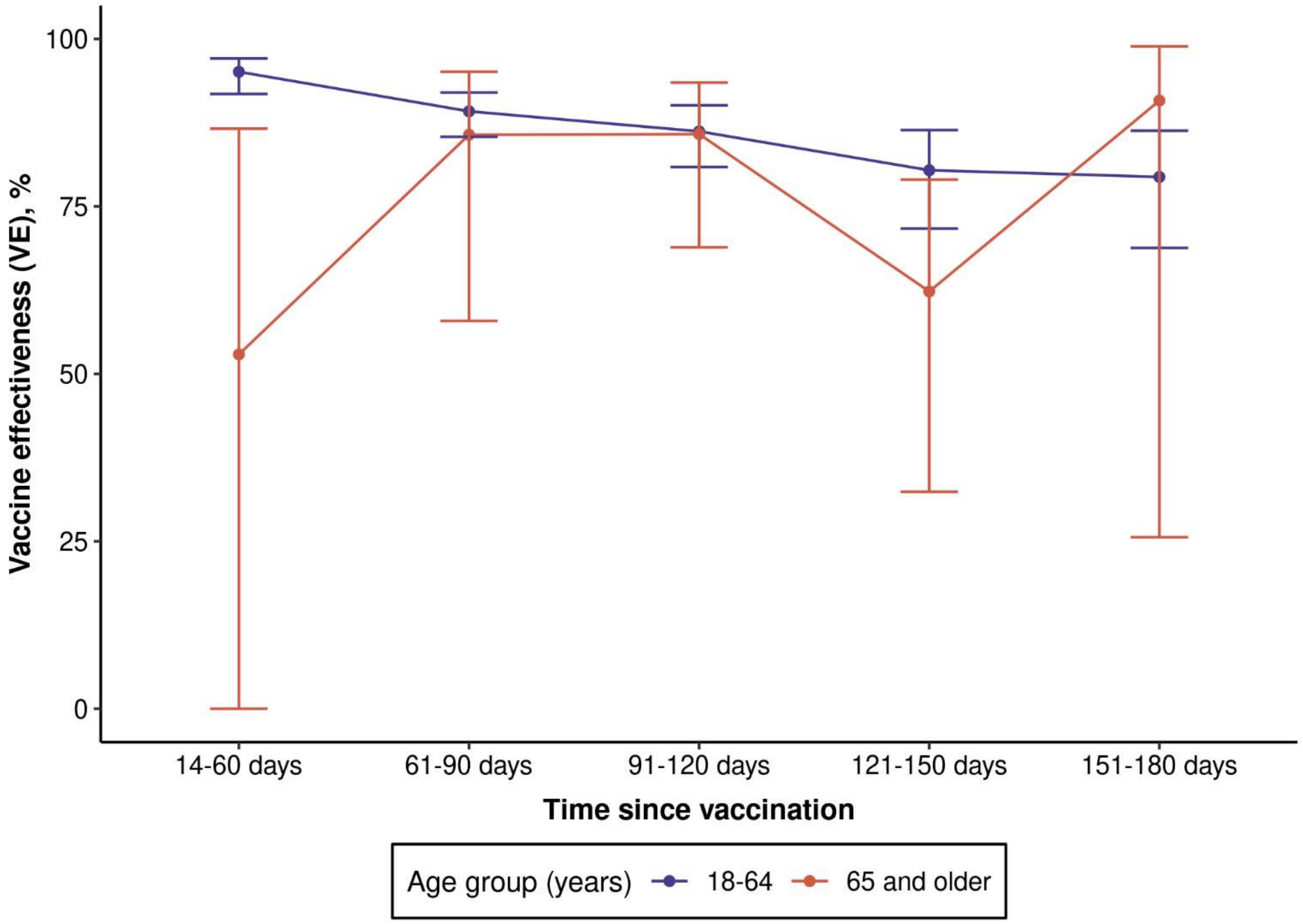

| | Test Positive | | Test Negative | | Adjusted[a] VE (95%CI) |
|---|---|---|---|---|---|
| | Vaccinated n (%) | Unvaccinated n (%) | Vaccinated n (%) | Unvaccinated n (%) | |
| Age 18-64 years | | | | | |
| Overall | 185 (9.7) | 1721 (90.3) | 4137 (43.4) | 5393 (56.6) | 87.9 (85.5, 89.9) |
| 14-60 days | 17 (0.9) | 1721 (90.3) | 948 (9.9) | 5393 (56.6) | 95.1 (91.8, 97.1) |
| 61-90 days | 53 (2.8) | 1721 (90.3) | 1397 (14.7) | 5393 (56.6) | 89.2 (85.4, 92.0) |
| 91-120 days | 46 (2.4) | 1721 (90.3) | 872 (9.2) | 5393 (56.6) | 86.2 (80.9, 90.1) |
| 121-150 days | 39 (2.0) | 1721 (90.3) | 536 (5.6) | 5393 (56.6) | 80.4 (71.7, 86.4) |
| 151-180 days | 30 (1.6) | 1721 (90.3) | 366 (3.8) | 5393 (56.6) | 79.4 (68.8, 86.3) |
| Age 65 years and older | | | | | |
| Overall | 47 (38.8) | 74 (61.2) | 451 (74.5) | 154 (25.5) | 75.2 (59.6, 84.8) |
| 14-60 days | 4 (3.3) | 74 (61.2) | 29 (4.8) | 154 (25.5) | 52.9 (0.0, 86.6) |
| 61-90 days | 5 (4.1) | 74 (61.2) | 68 (11.2) | 154 (25.5) | 85.7 (57.9, 95.1) |
| 91-120 days | 10 (8.3) | 74 (61.2) | 151 (25.0) | 154 (25.5) | 85.8 (68.9, 93.5) |
| 121-150 days | 27 (22.3) | 74 (61.2) | 174 (28.8) | 154 (25.5) | 62.3 (32.4, 79.0) |
| 151-180 days | 1 (0.8) | 74 (61.2) | 29 (4.8) | 154 (25.5) | 90.8 (25.6, 98.9) |

[a] Model adjusted for smoking, Charlson comorbidity score, frailty index, liver disease, pregnancy, history of COVID-19 diagnosis, number of outpatient and virtual visits, preventive care, medical center area, month of specimen collection, specimen type.
